# Genetic architecture of Multiple Sclerosis patients in the French national OFSEP-HD cohort

**DOI:** 10.1101/2025.04.28.25326562

**Authors:** Julien Paris, Nayane S.B. Silva, Igor Faddeenkov, Martin Morin, Léo Boussamet, Stanislas Demuth, Mitra Barzine, Anna Serova-Erard, François Cornelis, Sonia Bourguiba-Hachemi, Sophie Limou, Francis Guillemin, Sandra Vukusic, Romain Casey, Jonathan Epstein, Anne Kerbrat, Emmanuelle Leray, Eric Thouvenot, Guillaume Mathey, Laure Michel, Emmanuelle Le Page, Jerome De Seze, Christine Lebrun-Frenay, Caroline Papeix, Jonathan Ciron, Pierre Clavelou, Eric Berger, Aurelie Ruet, Thibault Moreau, Olivier Casez, Pierre Labauge, Abir Wahab, Gilles Defer, Amélie Dos Santos, Thomas David, Inès Doghri, Elisabeth Maillart, Laurent Magy, Helene Zephir, Olivier Heinzlef, Bertrand Fontaine, Laureline Berthelot, David-Axel Laplaud, Nicolas Vince, Pierre-Antoine Gourraud, OFSEP Biology group

**Affiliations:** Nantes Université, CHU Nantes, INSERM, Centre de recherche en Transplantation et Immunologie Translationnelle, UMR 1064, F-44000, Nantes, France; CHU de Strasbourg, Department of Neurology and Clinical Investigation Center, CIC 1434, INSERM 1434, F-67000 Strasbourg, France; CHU Clermont-Ferrand, Génétique – Oncogénétique Adulte – Prévention (GENOAP), Clermont-Ferrand; Université Clermont Auvergne, UFR de Médecine et des professions paramédicales, Clermont-Ferrand; CHRU, INSERM, Université de Lorraine, CIC Clinical Epidemiology, Nancy, Grand Est, France, Université de Lorraine, INSERM, INSPIIRE, Paris, Île-de-France, France; Hospices Civils de Lyon, Service de Neurologie, sclérose en plaques, pathologies de la myéline et neuro-inflammation, Bron, France, 69677; Observatoire Français de la Sclérose en Plaques, Centre de Recherche en Neurosciences de Lyon, INSERM 1028 et CNRS UMR 5292, Lyon, France, 69003; Université de Lyon, Université Claude Bernard Lyon 1, Lyon, France, 69000; Eugène Devic EDMUS Foundation Against Multiple Sclerosis (a government approved foundation) Bron, France, 69677; Neurology Department, CRC-SEP Rennes, Rennes Clinical Investigation Center CIC 1414, Rennes University Hospital Rennes University INSERM, CHU Pontchaillou, University, F-35000 Rennes France; Univ Rennes, EHESP, CNRS, INSERM, Arènes—UMR 6051, RSMS (Recherche sur les Services et Management en Santé)—U 1309, Rennes, France; CHU Nimes, Service de Neurologie, Univ Montpellier, Nimes, France; Institut de Génomique Fonctionnelle, Univ Montpellier, CNRS, INSERM, Montpellier, France; Nancy University Hospital, Department of Neurology, Nancy, France. Université de Lorraine, APEMAC, F-54000 Nancy, France; Neurology department; Clinical Neuroscience Centre, CIC_P1414 INSERM, Rennes, University Hospital, Rennes University; CHU Pontchaillou, CIC1414 INSERM, F-35000 Rennes France; Neurology, UR2CA_URRIS, Centre Hospitalier Universitaire Pasteur2, Université Nice Côte d’Azur, Nice, France; Foundation Adolphe de Rothschild Hospital, Department of Neurology, F-75000 Paris, France and université Paris -cité; CHU de Toulouse, CRC-SEP, department of Neurology, F-31059 Toulouse Cedex 9, France; Université Toulouse III, Infinity, INSERM UMR1291 - CNRS UMR5051, F-31024 Toulouse Cedex 3, France; CHU Clermont-Ferrand, CRC SEP Auvergne, Department of Neurology, and INSERM NeuroDol U1107, 63003 Clermont-Ferrand; CHU de Besançon, Service de Neurologie 25 030 Besançon, France; Department of Neurology, University Hospital of Bordeaux, 33076 Bordeaux, France; Neurocentre Magendie, Bordeaux University, INSERM U1215, 33000 Bordeaux, France; CHU de Dijon, Department of Neurology, EA4184, F-21000 Dijon, France; CHU Grenoble Alpes, Department of Neurology, Neurology MS Clinic Grenoble, Grenoble Alpes university hospital, Grenoble, F-38700 La Tronche; T-RAIG, TIMC-IMAG, Grenoble Alpes University, France; CHU de Montpellier, MS Unit, F-34295 Montpellier Cedex 5, France; University of Montpellier (MUSE), F-34000 Montpellier, France; APHP, Hôpital Henri Mondor, Department of neurology, F-94000 Créteil, France; CHU de Caen, MS expert centre, Department of Neurology, avenue de la Côte-de-Nacre, Normandy University, 14033 Caen, France; CHU La Milétrie, Hôpital Jean Bernard, Department of neurology et Centre d’Investigation Clinique, CIC INSERM 1402, F-86000 Poitiers, France; CHU de la Martinique, Department of Neurology, F-97200 Fort-de-France, France; CHU de Tours, Hôpital Bretonneau, CRC SEP and department of neurology, F-37000 Tours, France; Département de neurologie, Hôpital Pitié-Salpêtrière, APHP, Paris; Centre de Ressources et de Compétences SEP Paris, France; CHU de Limoges, Hôpital Dupuytren, Department of Neurology, F-87000 Limoges, France; CHU Lille, CRCSEP Lille, Univ Lille, U1172, F-59000 Lille, France; Hôpital de Poissy, Departement of Neurology, F-78300 Poissy, France; From the Sorbonne Université - Institut du Cerveau - Paris Brain Institute - ICM, Inserm, CNRS, APHP, Hôpital Pitié Salpétrière Univ. Hosp., DMU Neuroscience 6; Inst. of Cardiometabolism and Nutrition, Sorbonne-universités-Upmc 06, INSERM, CNRS; Laboratoire des Signaux et Systèmes (L2S), CNRS-CentraleSupélec, Université Paris-Saclay; Sorbonne Université (B.S.), Institut du Cerveau - Paris Brain Institute - ICM, Inserm, CNRS, APHP, Hôpital St. Antoine-HUEP; and INSERM (B.F.), SU, AP-HP, Centre de recherche en Myologie-UMR974 and Service of Neuro-Myology, Institute of Myology, University hospital Pitié-Salpêtriere.; Nantes Université, CHU Nantes, Pôle Hospitalo-Universitaire 11: Santé Publique, Clinique des données, INSERM CIC 1413, F-44000, Nantes

**Keywords:** Multiple sclerosis, France, OFSEP-HD, genetics, genomics, ancestry analysis, genomic proportions, bioinformatics, population genetics, Human Leukocyte Antigens, HLA haplotypes, HLA alleles

## Abstract

Multiple Sclerosis (MS) is a central nervous system (CNS) autoimmune inflammatory disease targeting the myelin sheath and affecting 2.8 million patients worldwide, mostly in economically advanced countries. The OFSEP-HD (French Multiple Sclerosis Registry - High Definition) multi-centric cohort comprises 2,667 genetic samples of patients with MS including 5 years of clinical, biological and imaging follow up. Here we described the genetic background of the cohort using data generated from the Affymetrix Precision Medicine Research Array (PMRA) genotyping chips to collect 888,799 genomic variants, and up to 8.5 million variants after imputation. Our analysis focused on genetic ancestry, admixture analysis and Human Leukocyte Antigen (HLA) including haplotypes inference. Principal Components Analysis (PCA) clustering identified seven ancestral clusters with 2177 patients (85.6 %) from clearly defined European ancestry. We observed 232 MS patients from North-African genetic ancestry while 120 of those patients (51.7%) did not self-report North-African origins, highlighting once again the limitations of self-assessed population descriptors. To promote data sharing we implemented the generation of a realistic and anonymous synthetic dataset using an adaptation of a known synthetic data generation methodology. This work unveils the genetic landscape and heterogeneous profiles of the OFSEP-HD cohort and proposes an open synthetic genetic dataset for further analyses.

## Introduction

Multiple Sclerosis (MS) is a complex autoimmune disorder characterized by the demyelination of nerve fibers in the central nervous system (CNS) yielding to heterogeneous symptomatology including cognitive dysfunction, muscle-related problems, mental health issues, vertigo, vision and sexual problems. The number of MS patients worldwide was estimated to 2.8 million with a prevalence of 35.9 per 100,000 inhabitants^1^. MS predominantly affects women, accounting for approximately 70% of cases demonstrating the significant gender disparity in susceptibility^1,2^. MS is highly prevalent in most economically advanced countries including France, with a prevalence of 197.6 per 100,000 inhabitants, which is more than five times greater than the worldwide average prevalence^3^. This highlights the substantial disease burden to the public health regarding healthcare quality and financial costs^4^.

MS pathogenesis is caused by the interplay between both environmental and genetical components. The well-known latitude gradient of MS demonstrates lowest prevalence regions near the equator and high prevalence regions outside of the tropics especially in North-Western countries^1^. A South-West to North-East increasing latitude gradient is also observable in metropolitan France, demonstrating the impact of the gradient even at a lower regional scale^3^. This gradient is thought to be primarily driven by the lack of sun exposure and vitamin D deficiency, more prominent as we get further from the equator^5^. Moreover, Epstein-Barr Virus (EBV) and CytoMegaloVirus (CMV) infections during lifetime, smoking and childhood obesity, contribute significantly to MS onset^6,7^. Nevertheless, this latitude gradient might also partly result in healthcare disparities across the world, with lowest prevalence regions bearing fewer data due to a deficit of neurologist and MS specialist, in turn yielding to a systemic bias towards fewer MS diagnostics in those regions^1^.

MS is a polygenic disease, resulting from both previously stated environmental factors and the cumulative effect of multiple genetic variations that influence susceptibility to the disease, as opposed to Mendelian diseases, which are mostly caused by a single genetic mutation. The most recent GWAS (Genome Wide Association Study) identified a total of 233 MS Single Nucleotide Polymorphism (SNP) associated to MS susceptibility, with 32 located in the Major Histocompatibility Complex (MHC) region explaining most of the phenotypic variance, and 1 located on the X chromosome^8^. Recent insights also suggest the role of genetics on disease severity and progression with one significant variant discovered to date^9^.

The French population’s genetic architecture stems from two major ancestries: early Neolithic farmers from Anatolia and steppe pastoralists from Eurasia. While Neolithic farmer DNA predominates in modern France, steppe pastoralist contribution is more pronounced in the north^10^. Over centuries, historical cultural and political borders have shaped the genetic architecture of today’s French population. Indeed, genetic clusters in France mostly align with neighboring countries, reflecting shared histories and interactions^10^. Moreover, France’s strategic location on Europe’s western edge has made it a key migration route between the continent’s south and north, enriching its genetic diversity. Notably, well-documented migration between North Africa and France, driven by historical, cultural, and economic ties, has contributed to human genetic diversity in southern Europe, including France.

However, studying genetic ancestry in France is historically sensitive, and faces legal constraints explaining the few genetic studies conducted on French population with genome wide data^10^. Additionally, most patients in genetic studies focusing on polygenic diseases like MS are from European genetic ancestry^13,14^. Nevertheless, initiatives like the French Multiple Sclerosis Registry – High Definition Cohort (OFSEP-HD) and its 2,667 samples with genetic data, is an opportunity to study MS genetics with high quality nation-wide data collection^15,16^. While the OFSEP-HD cohort is instrumental in numerous published MS studies, its genetic architecture has not been studied yet^17,18^. This study provides the first genetic analysis of the OFSEP-HD population, highlighting the role of ancestry and MS variants including MHC variations for future research into genetic factors influencing MS within the French population.

## Materials and methods

### Data collection

The OFSEP-HD observational study, totalizing 2,667 genetic samples of MS patients from the historical OFSEP study collected from various centers distributed across France, represents an ongoing cohort study initiated in 2018 with annual high quality follow-up^15,18^. All OFSEP-HD patients fulfilled MacDonald2017+ diagnosis criteria at inclusion^19,20^. Appropriate informed consent was obtained from all OFSEP-HD patients^20^. Data were generated by participating neurologists in the framework of the French MS registry (OFSEP)^15,16,18,20,21^. The cohort includes different types of demographic variables including patients’ and parents’ origins, sex, birthdate, Expanded-Disability-Status-Scale (EDSS) (scale measuring the level of disability), pregnancies, relapses, treatments and type of MS course at inclusion in addition to annual MRI data and sample biocollection^20,22,23^.

Genetic data were acquired through genotyping conducted at the Curie Institute’s Genotyping platform in Paris, utilizing the Affymetrix Precision Medicine Research Array (PMRA) chip, representing a total of 888,799 genotyped variants.

The OFSEP-HD cohort was compared to a subset of 2,405 reference individuals from the 1000 Genomes Project population, comprising individuals from East-Asian, South-Asian, American, European, and African ancestries^24^. In addition, a North African reference cohort of 154 non MS individuals from Arauna et al. was utilized to accurately characterize genetic ancestry in the light of its 486,252 genotyped variants^25,26^.

### Processing, imputation, and quality controls of genotyping data

Quality control (QC) procedures were applied to OFSEP-HD dataset and North-African reference dataset. No additional quality control or imputation was performed on the 1000 Genome reference dataset^27^. Technological QC on the OFSEP-HD dataset using Axiom Analysis Suite 2.0 selected 869,082 high-quality variants (97.8%).

A second QC filtering procedure was implemented using Python 3.11 with PLINK 1.9/2 and bcftools 1.13 following previously published guidelines^27^. Pre-imputation filters and thresholds applied to OFSEP-HD genetic samples are listed below:

1. Individual level and variant level missingness (>2%)
2. Sex-discrepancies (Male: F>0.8; Female: F≤0.4)
3. Exclusion of X/Y/mitochondrial chromosomes
4. Minor Allele Frequency (MAF<1%)
5. Hardy-Weinberg Equilibrium (HWE<1e^-10^)
6. Relatedness (p̂>0.2)

Processing steps of North-African reference dataset are detailed in Table S1 and a flowchart of the QC procedures in Figure S1^25^.

Pre-imputation quality controls of OFSEP-HD population resulted in 424,132 remaining SNPs and 2,542 patients. The filters removing most of the variants were the MAF filters and sexual and mitochondrial chromosome variants exclusion. Imputation of OFSEP-HD genetic data was then performed using Topmed Michigan Imputation Server to maximize overlap with reference datasets during ancestry analysis. Post-imputation QCs were also performed on OFSEP-HD dataset, excluding variants with insufficient imputation quality (r²<0.5) or low frequency (MAF<10%) for genetic ancestry analyses. Moreover, we filtered again for variants considerably deviating from HWE<1e^-10^.

### Ancestry analysis

We performed a Principal Component Analysis (PCA) of the 2,542 OFSEP-HD patients with all reference populations using the Eigensoft software. The analysis considered the twenty first principal components obtained from the 9,326 common independent SNPs to OFSEP-HD and reference datasets. A supervised K-Means clustering approach was then implemented to identify ancestry superpopulation clusters on the PCA (Figure S2)^28^.

### Demographic and clinical description: self-reported origins of OFSEP-HD patients

We analyzed demographic and clinical variables of the 2,542 remaining patients after quality controls and processing steps providing key insights into the composition of the cohort. We computed descriptive statistics to better understand the self-reported origins distribution of OFSEP-HD patients. Participants’ origins were reported via a Case Report Form (CRF) with three instances of the question formatted as follows: “Which geographical region are you originating from?”. Patients answered all questions, providing their own and their parents’ geographical origins.

### Reference populations selection and admixture proportions of European, African and East-Asian genome

Admixture analysis was performed using admixture 1.3.0 software. The reference parental populations for admixture analysis were selected from the 1,000 Genome dataset to include the three most discretizing ancestries: European, African, and East-Asian, thereby excluding North-African, South-Asian and American admixed ancestries^26,29,30^. More details are available in Supplemental Materials.

### Self-reported origins and genetic ancestry: a comparative statistical analysis

Patients’ and parents’ origins were compared to genetically inferred ancestral backgrounds. Self-reported origins of OFSEP-HD patients and their parents were collected through the previously mentioned CRF. Mapping between ancestry clusters and self-reported origins is shown in Table S2 and the methodology used to select clusters for statistical analyses is discussed in Figures S3-4 and Tables S3–4. Proportion of discordant pairs and Cohen’s kappa score were used as a measure of agreement between patients’ self-reported origins and genetic ancestry. Proportion of discordant pairs was calculated as 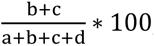, with *a*, *b*, *c* and *d* being respectively the upper left, upper right, lower left and lower right cells of a 2×2 contingency table. Margin of Errors (95%) (MoE_95%_) were computed as 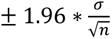 with *σ* the standard deviation and *n* the sample size^31^. Statistical significance of comparisons were assessed using two-sided statistical association tests from independent data arrangements^32^. We performed a quantitative statistical analysis comparing European and African genome proportions within genetic clusters regarding patients self-reported origins status.

### HLA imputation: alleles and haplotypes

Human Leukocytes Antigens (HLA) genotypes were imputed from genotyped SNPs through SNP-HLA Reference Consortium (SHLARC), a widely recognized and extensively used resource for imputing *HLA* alleles from SNP data, employing the HIBAG R package and a large multi-ethnic reference panel^33,34^. We also compared the proportion of patients with at least one *HLA-DRB1*15:01* most associated MS susceptibility risk allele between ancestry clusters using Mann-Whitney U test. Further, we performed individual-level haplotypes imputation using HLA2Haplo tool from Easy-HLA software suite independently for each ancestry cluster using corresponding reference panels^35^. This tool inferred the phase of imputed HLA alleles for each considered OFSEP-HD individual. Imputed haplotypes include loci *HLA-A, HLA-B, HLA-C, HLA-DRB1,* and *HLA-DQB1*.

### Polygenic Risk Scores computation

Polygenic Risk Scores (PRS) provides a summary metric measure of genetic susceptibility risk of an individual to a given genetic-related disease. Here we computed PRS designed specifically for MS, in the OFSEP-HD population^36,37^. We had 210 MS susceptibility variants (including 20 in the MHC region) out of a total of 233 known MS associated susceptibility variations available in our dataset after SNP imputation, filtering (MAF<1%) and *HLA* alleles imputation (Table S5)^8^.

We compared a log-additive PRS model to a non-weighted Sum of Risk Alleles Count (SRAC) approach using Pearson’s correlation coefficient r², using scores computed from the global OFSEP-HD population as well as from specific ancestry clusters^36–38^. Since MS susceptibility variants were mostly found in patients from European genetic ancestry, we only compared distributions of both models between European and North-African genetic ancestries using Mann-Whitney U tests^8,37,39^. Furthermore, we assessed the phenotypic variance explained by the 20 available MHC region genetic variations compared to the 190 non-MHC SNPs at the whole cohort level using Genome-wide Complex Trait Analysis (GCTA) software^40^.

### Synthetic data generation

To promote data sharing and reusability of sensitive genetic data, we provided an anonymous – synthetic – GDPR-enforced dataset as an open resource using an adaptation of an already existing “patient-centric” local modelling strategy^41^. The methodology focuses on addressing privacy risks like singling out, linkability, and inference using metrics such as Hidden Rate (HR), Local Cloaking (LC), and Distance To Closest synthetic record (DTC). Signal retention was evaluated using Hellinger distances, correlation differences, and Gower’s distance permutation test^42^. Optimal parameters were identified as k=2 and 20 principal components, with regularization for rare modalities ensuring respect of biological constraints. The privacy-fidelity balance was improved by generating three parallel synthetic datasets and averaging features patient-wise. Methodological details are available in Supplemental Materials and the synthetic dataset on GitHub (https://github.com/jp3142/OFSEP_HD_public_avatar_dataset).

## Results

### Ancestry analysis: Principal Components Analysis of OFSEP-HD MS patients

Figure 1 shows the Principal Component Analysis (PCA) of OFSEP-HD MS patients compared to the 2 reference datasets: 1,000 Genomes and North-African cohort^25^. It highlights the genetic diversity of the MS French population, with a specific focus on the first two principal components explaining respectively 49.1% and 29.9% of the variance (79%). The 10 most contributing SNPs for each principal component are listed in Table S6. The best clustering of the OFSEP-HD population was obtained using 7 distinct clusters.

**Figure 1:**
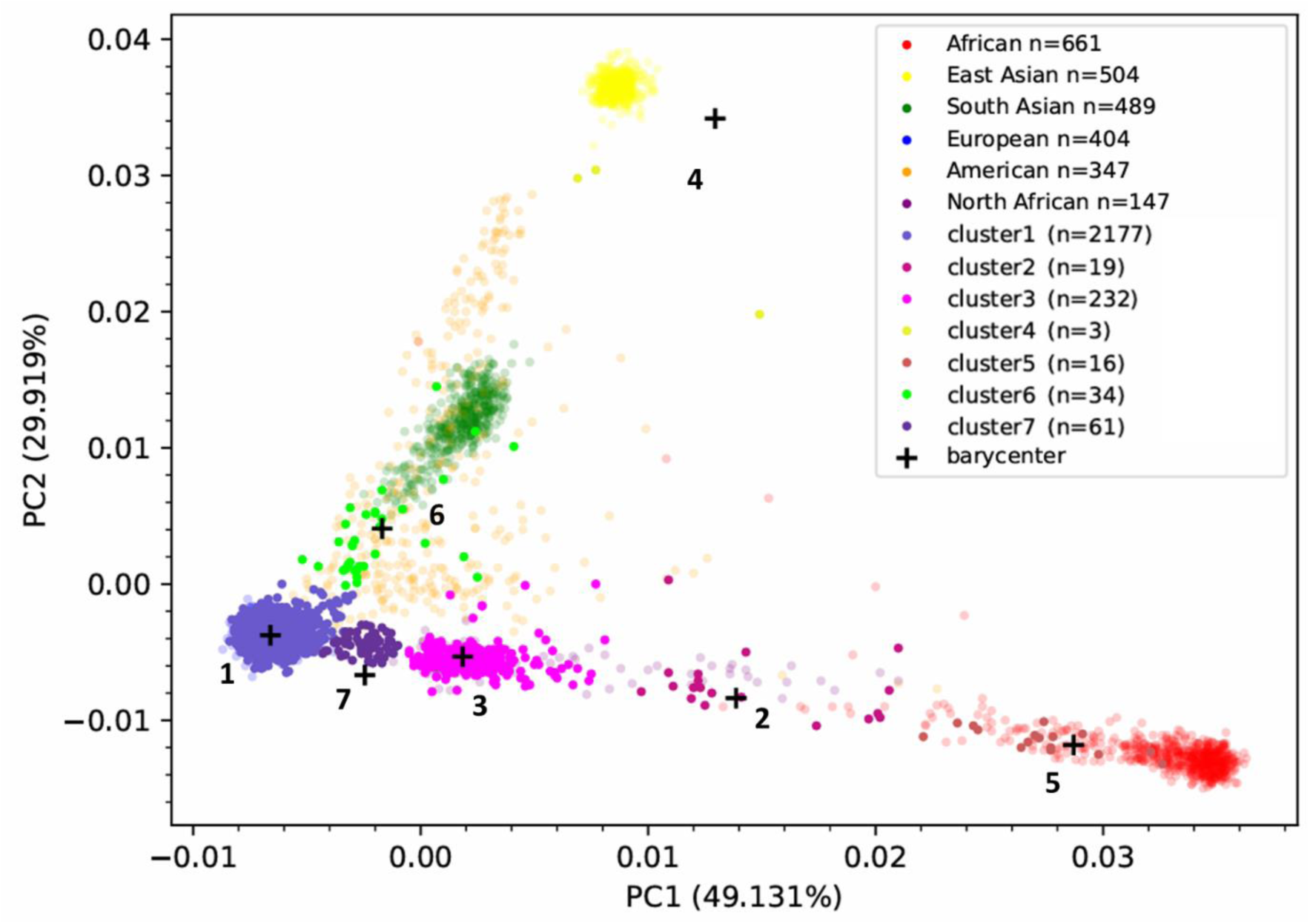
Principal Component Analysis (PCA) plot of the OFSEP-HD population based on the first two principal components (PC1 and PC2), overlaid with reference datasets. Each data point represents an individual. Semi-transparent data points represent reference individuals and opaque data points represent OFSEP-HD patients. Cluster 1: European. Cluster 2: Afro-Caribbean. Cluster 3: North-African. Cluster 4: East-Asian. Cluster 5: African. Cluster 6: South-Asian. Cluster 7: Admixed North-African and European.

Findings highlight a major proportion of patients with European genetic ancestry within the OFSEP-HD cohort, totalizing 2,238 MS patients (88%) assigned to clusters of European ancestry, clusters 1 (n=2,177) and 7 (n=61). Clusters 2 (n=19), 3 (n=232) and 5 (n=16) account for 267 MS patients (10.5%) with higher African ancestry. Clusters 4 (n=3) and 6 (n=34) totalize 37 MS patients (1.4%) with higher Asian ancestral background.

### Self-reported origins of OFSEP-HD patients

Table 1 shows 83% of patients who reported metropole France origins, while only 71% of fathers and 74% mothers self-reported such origins. Self-reported North-African origins account for 5% of patients and around 10% of their parents. About 6% of patients reported other European origins. The least represented origins are French overseas territories, Asian and Sub-Saharan African origins. Ninety-eight patients did not answer the question or did not know about their origins, and 15 patients reported to be originating from other parts of the world. Details about other demographic and clinical data are shown in Tables S7–9.

**Table 1:** Self-reported origins of 2,542 remaining OFSEP-HD patients after quality controls and processing steps. Count variables correspond to number of patients, and the % variables correspond to proportion in the OFSEP-HD population subset.

|  | Origin count | Origin % | Father's origin<br>count | Father's origin % | Mother's origin<br>count | Mother's<br>origin % |
| --- | --- | --- | --- | --- | --- | --- |
| **France metropole** | 2,110 | 83.01 | 1,812 | 71.28 | 1,889 | 74.31 |
| **French Overseas**<br>**Territories** | 14 | 0.55 | 19 | 0.75 | 16 | 0.63 |
| **Europe (besides**<br>**France)** | 156 | 6.14 | 238 | 9.36 | 231 | 9.09 |
| **North-Africa** | 127 | 5.00 | 272 | 10.70 | 233 | 9.17 |
| **Sub-Saharan**<br>**African** | 10 | 0.39 | 23 | 0.90 | 20 | 0.79 |
| **Asia** | 12 | 0.47 | 24 | 0.94 | 22 | 0.87 |
| **Others** | 15 | 0.59 | 27 | 1.06 | 30 | 1.18 |
| **Unknown** | 98 | 3.86 | 127 | 5.00 | 101 | 3.97 |

### Admixture proportions of European, African and East-Asian genome in OFSEP-HD cohort

We observed a major European genome proportion in the OFSEP-HD population with an average above 93.7% (Figure 2). There is also a substantial number of OFSEP-HD patients (12.3 %) exhibiting more than 10% of African genome (Figure 2 and Table S10). Moreover, most MS patients showing significant African genome ancestry are estimated as part of North-African (cluster 3; n=232) genetic cluster. Distributions of genome proportions among cluster 3 is composed in majority by European genetic components (75.5% in average) compared to the African genome proportions ranging from 16.7% to 38.9%. Moreover, we found a non-neglectable East-Asian genome proportion for some MS patients, with 101 (<4%) with at least 5% of East-Asian genome ancestry and 2 patients with more than 85% of East-Asian ancestry (Figure 2, Table S10).

**Figure 2:**
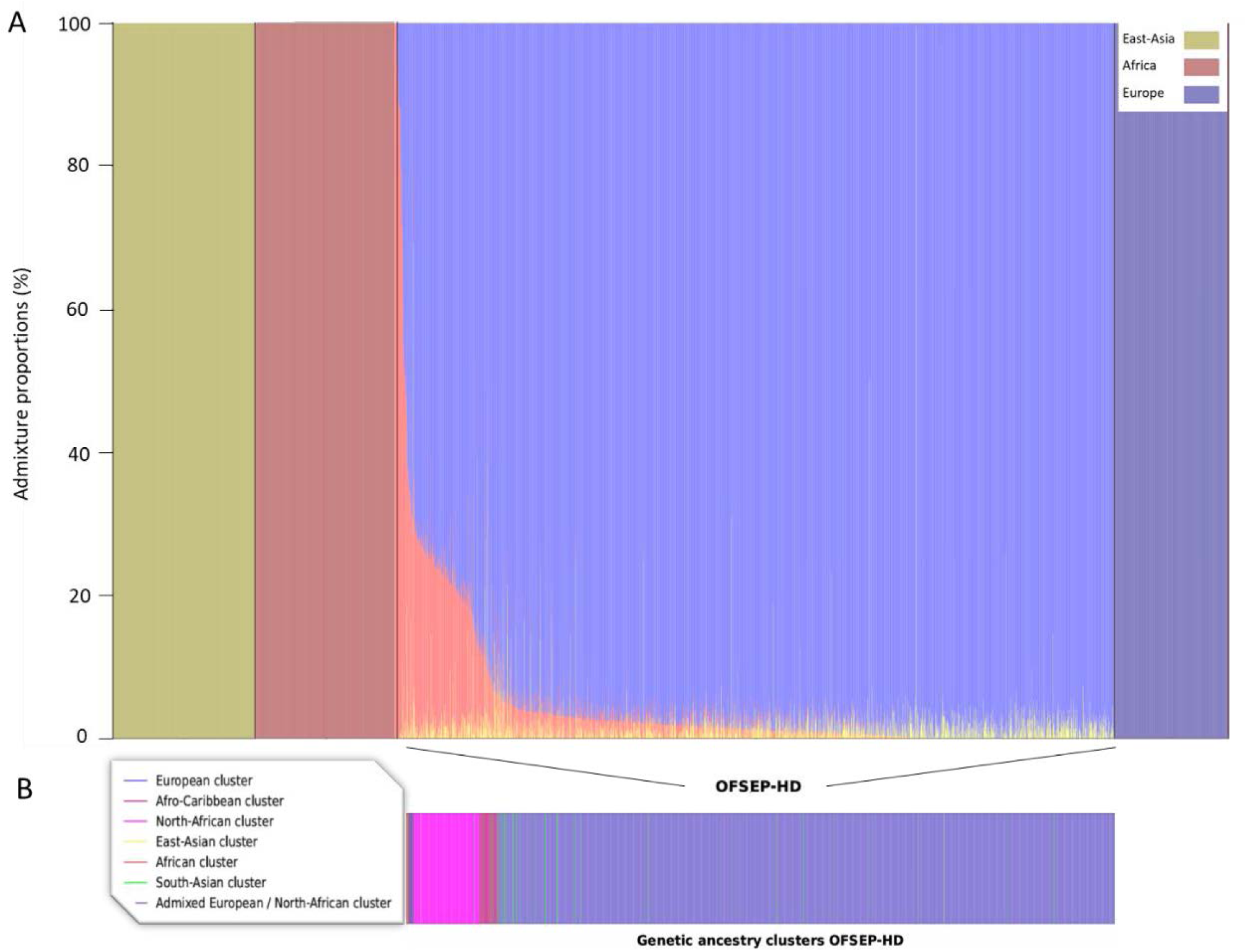
(A) Admixture plot showing whole-genome admixture proportions of African (red), East Asian (yellow), and European (blue) ancestry for each of the 2,542 OFSEP-HD patients, sorted by African genome proportion. Reference parental populations are shown on both sides of the OFSEP-HD section. (B) Mapping of genetic ancestry clusters previously identified using PCA for each patient.

### Self-reported origins and African-related genetic ancestry

We focused on qualitative analysis of North-African origins as it represents the second largest genetic group in the OFSEP-HD cohort, after European ancestry. Table 2 illustrates the distribution of OFSEP-HD patients based on their North-African self-reported origins and genetic ancestry statuses.

**Table 2:** Distribution of OFSEP-HD patients regarding North-African self-reported origins and genetic ancestry.

|  | North-African<br>genetic ancestry | Other genetic<br>ancestry | Total |
| --- | --- | --- | --- |
| North-African self-reported origins | 112 | 15 | 127 |
| Other self-reported origins | 120 | 2295 | 2415 |
| Total | 232 | 2310 | 2542 |

Cohen’s Kappa metric (κ = 0.598±0.001) and the low proportion of discordant pairs (5.31%) illustrate a moderate to substantial agreement between self-reported North-African origins and genetic North-African ancestry (Table 2). Remarkably, among 127 patients self-reporting North-African origins, 15 patients (11.8%) have a genome that does not reveal marked North-African genetic background. Moreover, 51.7% of patients from North-African genetic ancestry self-identified as from other origin. Further analyses about North-African patients are presented in Figure S5. Likewise, we detected a strong underestimation of self-reported Sub-Saharan-African (p<0.001) and Afro-Caribbean origins (p<0.05) compared to their respective PCA-defined ancestry clusters (Figures S6–10; Tables S11–14).

### North-African patients and admixed genome proportions

Among non-North African genetic ancestry patients, no significant differences in African and European genomic components were found between those self-reporting North African origins and those who did not (Table S15), confirming consistency between PCA-defined ancestry clusters and ancestral genomic composition. Thus, we focus on non-North African genetic ancestry patients.

We found that self-reported North-African patients from other genetic ancestry have a statistically significant higher proportion (Mann-Whitney U: p<1e-4) of African genome compared to patients who did not report such origins (Table S15; Figure S15). Nevertheless, the high variability associated to African genome proportions (CI_95%_ [3.01%; 28.65%]) and consequently European genome proportions (CI_95%_ [70.44%; 95.78%]; Table S16) of self-reported North-African patients from other genetic ancestry demonstrates the admixed nature of this subpopulation. Further details on performed analyses and European genome proportions of OFSEP-HD North-African MS patients are discussed in Figures S11–14 and S16-19. Details and analyses of other genetic ancestry clusters and origins are available in Tables S17–28.

### Parents’ origins: a better proxy of patients’ genetic ancestry?

We detected a significant association between parents’ origins and children estimated genetic ancestry (p<1e-4). However, we found no significant statistical association (p=0.77) between parents’ North-African origins and their child’s genetic ancestry when only one parent reported North-African origins, demonstrated by high discordance for both fathers (84.1%) and mothers (86.7%) (Figure 3A and 3B; Table S29). Nevertheless, we observed a substantial agreement with patients’ North-African genetic ancestry when both parents have common origins (84.7%, Figure 3C) also highlighted by a higher Cohen’s Kappa value (κ=0.64), and a lower proportion of discordant pairs (3.11%) (Tables S30–33), compared to what was obtained previously with patients’ self-reported origins (Table 2). Additionally, 74.1% of MS patients with North-African genetic ancestry reported both parents as from North-African origins (Figure 3D). Analysis of other parents’ origins and genetic ancestries are available in Tables S34–37.

**Figure 3:**
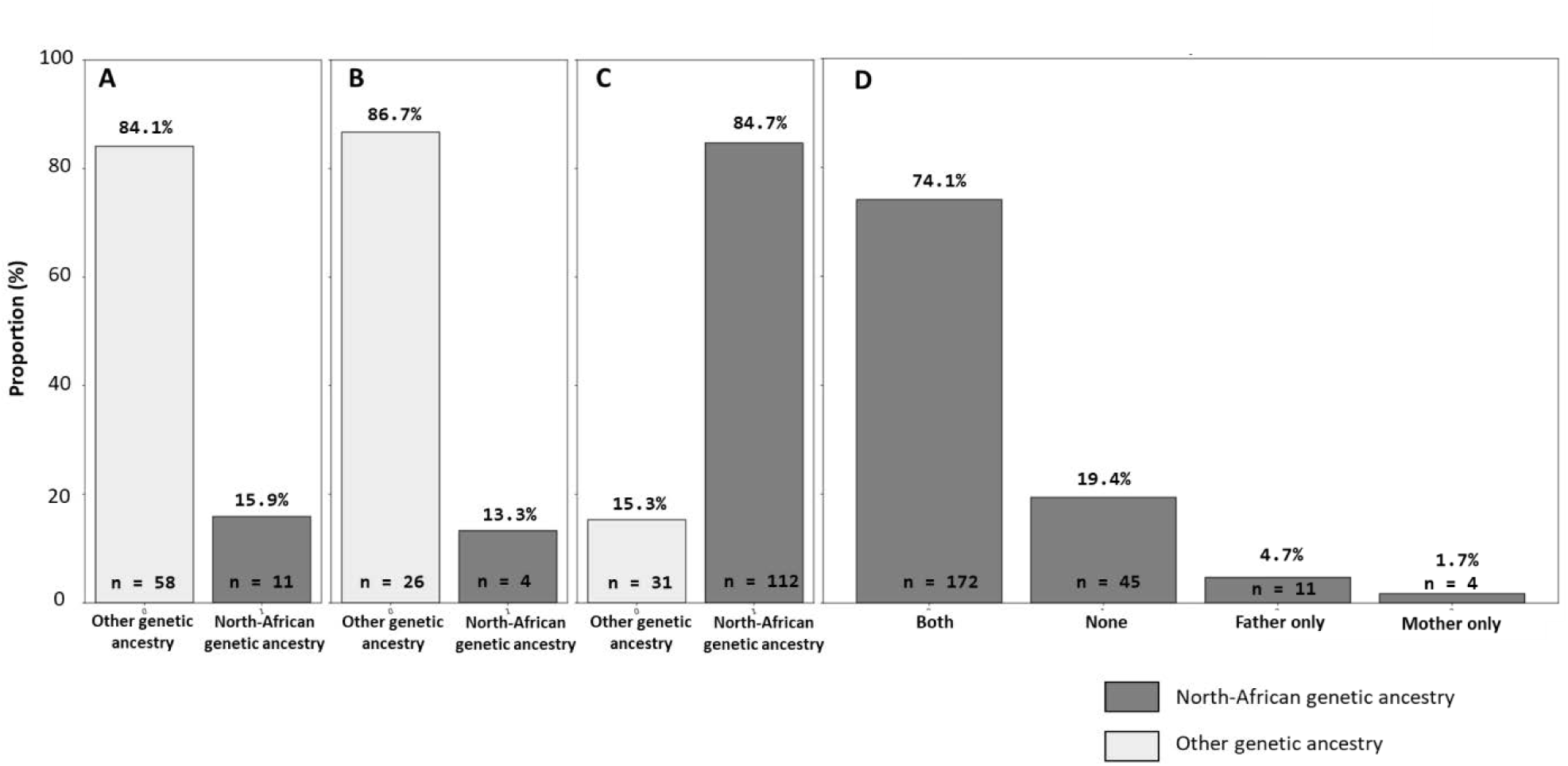
(A) North-African genetic ancestry status among patients with only the father reported as from North-African origins. (B) North-African genetic ancestry status among patients with only the mother reported as from North-African origins. (C) North-African genetic ancestry status among patients with both parents reported as from North-African origins. (D) Parents’ North-African origins status of patients from North-African genetic ancestry (Both: both parents were reported as from North-African origins, None: no parents were reported as from North-African origins, Father only: the father is the only parent reported as from North-African origins, Mother only: the mother is the only parent reported as from North-African origins).

### HLA alleles imputation

*HLA* alleles imputation was performed on classical *HLA* genes *HLA-A, HLA-B, HLA-C, HLA-DPA1, HLA-DPB1, HLA-DQA1, HLA-DQB1* and *HLA-DRB1* and is particularly accurate for all genes. The average imputation post-probabilities ranging from 86.5% for *HLA-DRB1* to 98.9% for *HLA-DPA1*, indicate trustful results across all considered *HLA* class I and II genes. In addition, we found a significantly higher proportion of patients (p<1e-5) with at least one *HLA-DRB1*15:01* allele in the European genetic cluster (48.8%) compared to the proportion among the North-African genetic cluster (33.2%)^43^. Additional details about imputation quality distribution statistics are shown in Figure S20 and Table S38.

### HLA haplotypes imputation

*HLA* haplotypes imputation from imputed *HLA* alleles *HLA-A, HLA-B, HLA-C, HLA-DRB1* and *HLA-DQB1* achieved very high accuracy with an average post-probability of 82.9% across all haplotypes collectively. Figure S21 and Table S39 show distribution of post-probabilities by HLA locus. We summarize HLA diversity of the OFSEP-HD MS cohort in Table 3.

**Table 3:**
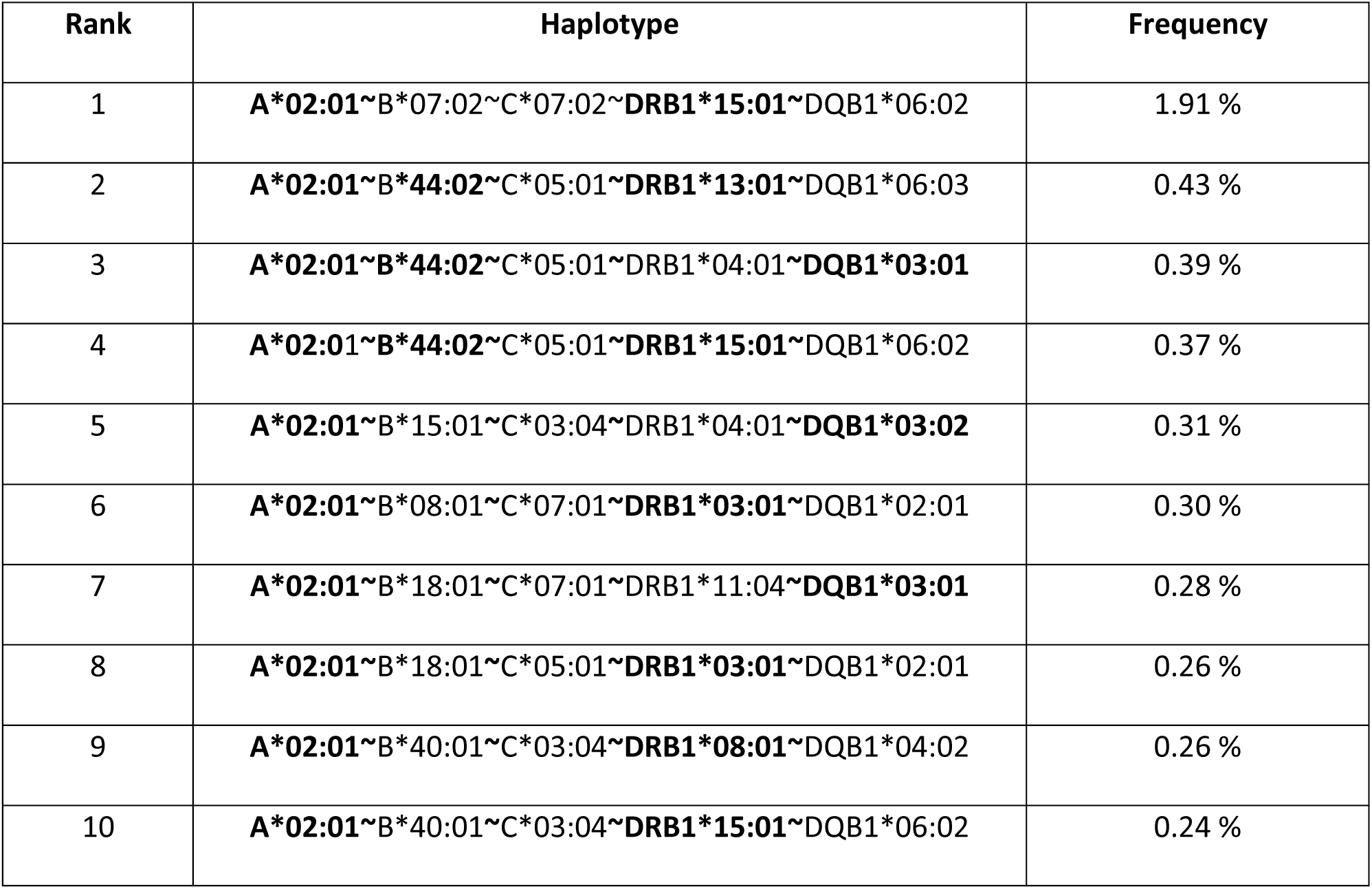
Description of the 10 most frequent imputed haplotypes including at least one MS associated HLA allele within the OFSEP-HD cohort. Bold alleles within haplotypes represent known MS associated HLA alleles^8,44^.

All of the ten most frequent HLA haplotypes including at least one HLA MS associated allele present the *HLA-A*02:01* protective allele, but some also contain *HLA-DRB1*15:01* risk allele. The most frequent HLA haplotype is ***A*02:01*∼***B*07:02***∼***C*07:02***∼*DRB1*15:01*∼***DQB1*06:02* (1.91%).

### Computing polygenic risk scores of French MS populations

We found an average log-additive score of 20.29±0.03 for 190 non-MHC SNPs, representing 201 risk alleles per patient. Including the 20 available MHC variations increased the score to 23.38±0.05 for 214 risk alleles in average. The log-additive model and SRAC showed high correlation (r²=0.91) for non-MHC SNPs, but correlation decreased substantially (r²=0.54) when MHC variants were included (Figure S22). Moreover, MHC variants increased the log-additive score by 15.23% while only increasing the average number of risk alleles by 6.46%, highlighting their larger effect on MS susceptibility. PRS distributions differed significantly between MS patients from North-African and European ancestries for all PRS models (p<1e-6). PRS scores distribution statistic details are available in Table S40 and Figure S23.

### Open-access synthetic data generation: a balance between privacy and fidelity

Using our synthetic data generation process, we created a highly anonymized dataset as indicated by key metrics following recommended thresholds: HR=99.49% (>90%); median LC=50 (>5), with DTC=1.8 (>0.2) and CDR=0.99 (>0.3)^41^. Signal retention metrics also show high synthetic dataset fidelity to original data with Hellinger distance distributions difference of 0.04 (<0.1) and distributions correlation difference of 1.92% (<10%). Univariate statistical analysis of generated and original datasets indicated four features (rs58394161, rs11809700, rs11578655, rs10801908) out of the 82 generated features where distributions in synthetic data were significantly deviating from original data distributions after Bonferroni correction for multiple testing (p<0.05). Gower’s distance permutation test showed no statistically significant overall differences from what would be expected by chance between generated and original datasets (p=0.38)^45^. Figure S24 shows datasets overlap on the two first dimensions of the reduced original Euclidean space.

## Discussion

The OFSEP-HD study provides valuable insights into the genetic landscape of the French MS population, revealing significant variations of ancestral origins of MS patients in France. We highlighted a significant proportion (232 patients; 9.1%) of MS French patients from North-African genetic ancestry, illustrating the substantial North-African ancestral background of the French population.

Regarding PCA ancestry analysis, we believe the proportion of variance explained by the two first components (79%) is adequate to reveal the key structure of the OFSEP-HD cohort genetic background. Cluster 7, an admixed North-African–European population, was treated as part of the European cluster in statistical analyses due to its closer proximity to the European reference population and lower average African genome proportion compared to the North-African cluster (Tables S3–4, Figures S3–4).

The study reveals differences between self-reported origins and genetic ancestry of French North-African MS patients. Only half North-African genetic ancestry patient self-identified as such, highlighting the significance of cultural affiliation over ancestral genetic backgrounds for patients born in France. Approximately 10% of these patients reported “unknown” or “other” origins, suggesting complexities in identity and societal perceptions within multicultural societies (Figure S10)^46^.

The phrasing of the OFSEP-HD survey question, “From which geographical region are you originating from?”, may lead to diverse interpretations, such as birthplace or upbringing^47^. Other biases, like form completion errors or practitioner misinterpretations, should also be considered^48^. Using the example of the MS French population from North-African origins, we highlight the misuse of self-reported origins as population descriptors, which can perpetuate inaccurate genetic ancestry presuppositions. We recommend against using such descriptors alone as genetic ancestry proxies. However, despite the contentious nature of geographical origins and ethnicity as population descriptors, we suggest not fully dismissing them in genetics and genomics research, as they provide insights to better characterize groups experiencing healthcare disparities, including genomic healthcare access^49^.

Our findings indicate a stronger alignment between reported North-African origins and genetic ancestry when both parents share the same origins. Discrepancies occur with differing parental origins. Caution is advised when using parental origins as a genetic ancestry proxy, as it may not fully represent the child’s genetic background, particularly with only one North-African parent. Among other genetic clusters, self-reported North-African origins show a significantly higher (p<1e-4) African genomic ancestry proportion (15.83%±12.82%) compared to other origins (2.54%±0.36%). However, the high variability (MoE95%=12.82%) suggests diverse reasons for self-reporting North-African origins.

Log-additive models for PRS calculation offer no significant advantage over the simpler SRAC approach for non-MHC autosomal SNPs, due to similar effect sizes and nearly identical impacts on PRS (0R≈1; Figure S22A)^8,44^. A log-additive approach is more suitable for MHC variants with larger effect sizes, unlike SRAC where all variants have equal impact (Figure S22B). Most MS variants, identified in European populations, may not fully capture genetic risks for other ancestries^37,39^. Our findings reveal significant PRS score differences and significantly lower *HLA-DRB1*15:01* allele proportions in patients from North-African genetic ancestry, suggesting reduced MS susceptibility. This must be contextualized within European-focused research, healthcare disparities, and environmental factors like vitamin D, contributing to a South-to-North prevalence gradient^3,50^. Current PRS may perpetuate healthcare disparities, emphasizing the need for more inclusive models^51^.

Finally, our adapted methodology of the Avatar algorithm significantly improved fidelity and privacy compared to single-generation synthetic avatar data^41^. Averaging parallel synthetic data generation in a patient-wise manner enhanced fidelity and privacy by combining multiple synthetic patients for each original patient. All privacy and signal metrics significantly exceeded standard thresholds, demonstrating the methodology’s flexibility in producing high-fidelity synthetic datasets compliant with GDPR^41^. This makes it a viable tool for future research. However, our study only performed univariate comparisons between original and synthetic datasets. Future work should verify the preservation of multivariate dependencies to maintain signal integrity.

## Conclusion

In this study, we characterized the genetic landscape of the French OFSEP-HD MS cohort into seven genetic ancestral clusters. We found that self-reported origins significantly underestimate North-African ancestry, highlighting their limitations as proxies for genetic ancestry; parental origins proved more reliable. In addition, French MS patients from North-African genetic ancestry exhibited admixture profiles closer to European than to sub-Saharan African populations. We also imputed HLA alleles/haplotypes and computed MS PRS. However, our findings demonstrate that current MS susceptibility variants are ill-suited for calculating PRS in non-European ancestries. This work establishes the first comprehensive framework for analyzing genetic diversity in the French OFSEP-HD cohort and propose a first open synthetic and GDPR-enforced genetic dataset for this population, including re-identification risk assessment. This advances synthetic data generation methodologies and paves the way for new methods tailored to genetic data’s sensitivity and complexity. Overall, it establishes a foundation for future MS research in France, addressing the national context of limited genetic ancestral studies due to cultural and ethical considerations.

## Data Availability

All data produced are available online at https://github.com/jp3142

## Acknowledgments

Data collection has been supported by a grant provided by the French State and handled by the “Agence Nationale de la Recherche,” within the framework of the “France 2030” programme, under the reference ANR-10-COHO-002” OFSEP; and the support of the “Eugène Devic EDMUS Foundation against multiple sclerosis”; as well as the contribution of the “ARSEP Foundation” and the contribution of the Biological Resources Centres (BRC) that provided biological sample. In addition, we thank the historical contributors of REFGENSEP, Lena Guillot-Noël and Isabelle Rebeix.

This work was also supported by the PRIMUS project (Projection in Multiple Sclerosis), part of a government grant managed by the French National Research Agency (Agence Nationale de la Recherche, ANR) as its 3^rd^ PIA, integrated into the “France 2030” plan under reference [ANR-21-RHUS-0014]. This work was supported by Nantes Métropole, Région des Pays de la Loire and European Union (FEDER) via the Programme d’investissements d’Avenir (NExT, SHLARC Project, Nantes Université).

Moreover, insights and encouragement from following collaborators were instrumental in the completion of this work. I am grateful to CR2TI for providing the necessary facilities and technical support, and Dr. Vincent Mauduit for his assistance with data analysis.

We thank the Curie genomic platforms for technical support and GWAS genotyping. We are also grateful to the Bioinformatics Core Facility BiRD, member of Biogenouest and Institut Français de Bioinformatique (IFB) (ANR-11-INBS-0013) for the use of their resources and their technical support.

## Authors’ contribution

**Data Curation**: M.M. performed the initial quality control using the Axiom Analysis Tool from Thermofisher, following standardized protocols. J.P. curated the final dataset from the original data.

**Formal Analysis**: N.S.B.S. conducted the HLA imputation using the HIBAG R tool and data reference from SNP-HLA Reference Consortium. L.B imputed HLA haplotypes and phase using HLA2Haplo interface.

**Investigation** J.P performed all other analyses and experiments related to the study.

**Data Anonymization and Synthetic Data Generation**: J.P. generated anonymized synthetic data using the Avatarization method from Octopize in the purpose of data sharing.

**Conceptualization**: J.P. with input from P.A.G. and N.V, as well as M.M and I.F.

**Supervision**: P.A.G., N.V.

**Writing – Original Draft:** J.P.

**Writing – Review & Editing**: P.A.G. and N.V. as well as I.F, L.B, and S.D contributed to discussions and validated the final manuscript.

### Web ressources

Bcftools 1.13: https://samtools.github.io/bcftools/

Plink 1.9 and 2: https://www.cog-genomics.org/plink/

Admixture 1.3.0: https://dalexander.github.io/admixture/

TopMed Michigan Imputation Server: https://imputation.biodatacataslyst.nhlbi.nih.gov/

Eigensoft - smartpca: https://github.com/chrchang/eigensoft

EasyHLA : https://easyhla.univ-nantes.fr/

HLA2Haplo (EasyHLA feature): https://easyhla.univ-nantes.fr/prediction/prediction.php

Avatar: https://github.com/octopize/avatar-python

Genome-wide Complex Trait Analysis (GCTA) 1v.94.1 Linux: https://yanglab.westlake.edu.cn/software/gcta/#Overview

## Data and code availability

Avatar dataset and pipeline scripts are available on github (https://github.com/jp3142).

## Declaration of generative AI in scientific writing

During the preparation of this work the authors used OpenAI’s GPT-4, as well as Mistral’s model “LeChat” in order to assist with language refining and rephrasing of certain sections. After using this tool/service, the authors reviewed and edited the content as needed and take full responsibility for the content of the publication.

