## supplemental_materials for "Genetic architecture of Multiple Sclerosis patients in the French national OFSEP-HD cohort" (from another version of this preprint: version 3, 2026-02-13)

#### Details on pre-imputation processing and quality controls of genotyping data

Original OFSEP-HD genetic data were collected in GRCh37/hg19 genome build version. The 1000Genomes genetic data were collected in GRCh38/hg38 genome build version and no additional quality controls were performed. We performed technological quality control (QC) procedure on OFSEP-HD data using a DishQC > 0.82 ensuring proper separation of AT and GC fluorescence signals from noise in nonpolymorphic test probes, and a probe call rate by sample of > 97%. Similarly, we applied a call rate of > 98.5% and > 95% for plates and genetic variants, respectively 869,082 variants (97.8 %) and 2,667 genetic samples (99.3 %) passed those technological QC steps.

The sequence of filtering steps is available in Table 1 for OFSEP-HD patients and in Table S1 for North-African reference individuals[1]. First, we performed the lift-over of North-African GRCh37/hg19 reference population to GRCh38/hg38 genome build version using a GRCh38/hg38 reference sequence, to align each dataset on the same genome reference build. This lift-over step leaded to the exclusion of 138 variants, yielding to 486,114 remaining variants. We then performed the removal of individuals and variants with a significant missingness level above the classical 2% threshold. We checked for sex discrepancies within the OFSEP-HD population, that might imply sample mix-ups, and we imputed missing sex data when needed. The F statistic corresponds to the X chromosome homozygosity estimate and is used to impute sex, usually with parameter F > 0.8 for males and F < 0.2 for female. However, in order to keep as many individuals as possible, we chose to impute sex using less stringent F statistic thresholds with a F >= 0.8 for males and F <= 0.4 for females. Indeed, some individuals self-reported as female had a F statistic between 0.2 and 0.4. Additionally, we excluded sexual chromosomes from the OFSEP-HD dataset and we removed Single-Nucleotide-Variants (SNV), with a Minor Allele Frequency (MAF) lower than 1% in both North-African reference and OFSEP-HD datasets. We then removed variants deviating from Hardy-Weinberg Equilibrium (HWE) with a filter threshold of $1x10^{-10}$ for OFSEP-HD and North-African reference data. Finally, we pruned the dataset to keep only independent SNP and removed any significantly related individuals and patients with a threshold $\hat{p} > 0.2$ [2], e.g. at least second degree relatives. Nevertheless, we did not perform SNP imputation on North-African or 1000 Genome reference data to retain only genotyped variants, more reliable for populations serving as reference. We filtered out variants with a MAF < 10% for our ancestry analyses. Finally, we excluded all SNPs with highly deviating frequencies (difference > 10%) from North-African and 1000 Genomes data, which may indicate low data quality.

| Order | Filter / preprocessing step | Exclusion threshold |
|---|---|---|
| **1** | Lift-over | / |
| **2** | Individual level and variant level missingness | > 2% |
| **3** | Minor Allele Frequency (MAF) | < 1% |
| **4** | Hardy-Weinberg Equilibrium (HWE) | $< 1e^{-10}$ |
| **5** | Relatedness | $\hat{p} > 0.2$ |
| **6** | Minor Allele Frequency (MAF) | < 10% |
| **7** | Overlapping pruned SNPs to all datasets | / |
| **8** | Highly deviating SNPs frequencies from 1000 Genome dataset | > 10% |
| **9** | AT/GC SNPs removal | / |

***Table S1****: Sequence of quality control filters applied to North-African reference genotyping data from Arauna et al. study*[1]*. All processing steps required for ancestry analysis are displayed here since no imputation was performed on reference datasets. We did not have access to sexual chromosomes for North-African individuals.*

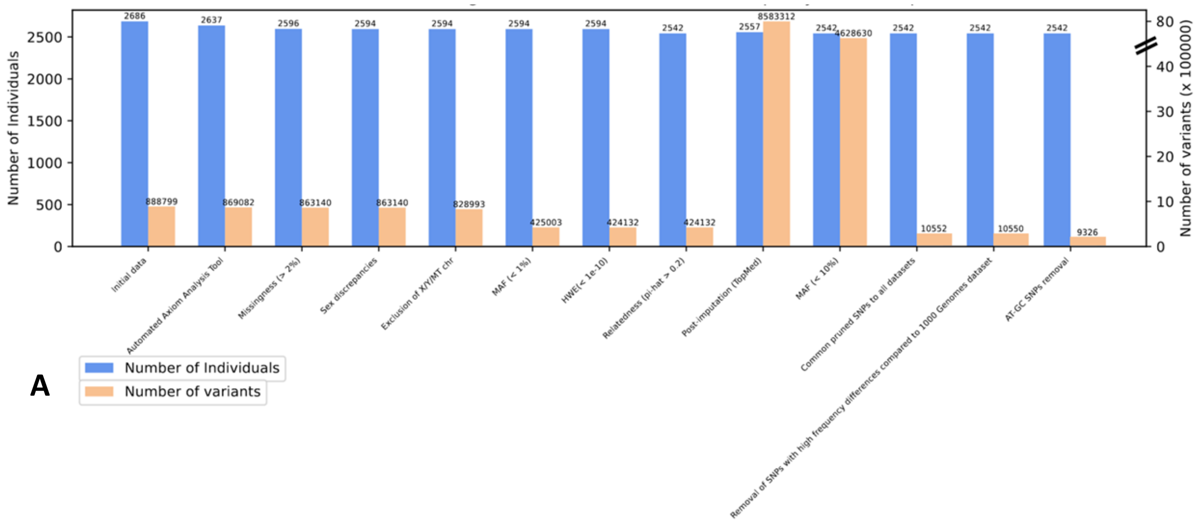


**A**

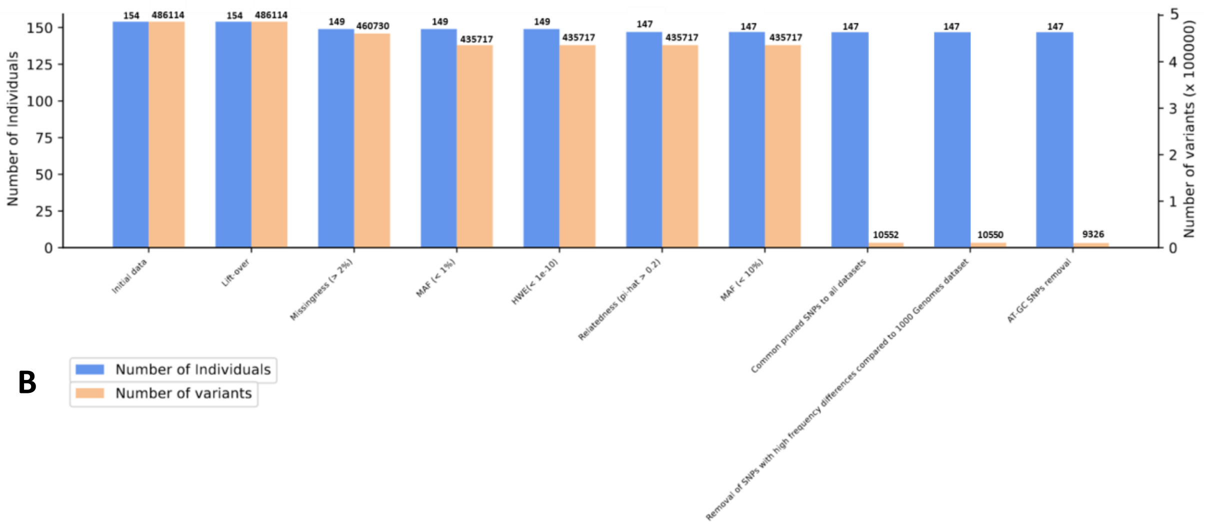


***Figure S1:*** *(A) Flowchart showing number of remaining variants and patients after each filter applied during pre and post imputation quality control filtering for OFSEP-HD population. (B) Flowchart of the number of remaining variants and individuals after each filter applied during quality control filtering for North-African reference population.*

Quality control procedure was performed independently for OFSEP-HD patients and North-African reference data, ensuring data maintains the same high standards of quality and reliability. Quality control steps resulted 2,542 patients and 424,132 SNPs for the OFSEP-HD cohort and 147 individuals and 435,717 SNPs for North-African reference datasets (Figure S1). Additionally, MAF (< 1%) and missingness filters (> 2 %) are the most constraining filters for North-African dataset, even though they are reducing the number of SNPs of only about 10.5%, probably because the dataset was preprocessed as part of the original study[1]**.**

### Details on SNP imputation and post-imputation processing of OFSEP-HD genotyping data

We used TOPMed reference panel “TOPMed r3" for imputation and we set $r^2$ filter to 0.3 to minimize file size by excluding all imputed SNPs with a $r^2$-value lower than 0.3. This $r^2$ metric represents the imputation quality computed as the estimated value of the squared correlation between imputed genotypes and true, unobserved genotypes. During post-imputation processing and quality controls, we excluded variants with insufficient imputation quality ($r^2$ <= 0.5) and a MAF lower than 10% corresponding to a common threshold used for ancestry analyses. We chose a relaxed $r^2$ filter of 0.5 to balance data quantity and quality, keeping as much variants as possible. However, we did not apply the MAF > 10% filter on North-African and 1000Genomes reference datasets to maximize overlap with filtered OFSEP-HD dataset. Moreover, we filtered for SNP deviating from HWE ($1e^{-10}$). We fixed the reference of OFSEP-HD dataset after imputation using the same reference sequence as the one used on North-African and 1000 Genome reference datasets.

**Details on ancestry analysis**

We discarded 99 Finnish individuals from 1000 Genome population from the analysis due to their distinct genetic configuration from the rest of the European population[3]. To set the number of ancestry clusters from the OFSEP-HD population, i.e., “k” parameter of the K Means algorithm, we implemented the “elbow” method, a statistical technique for identifying the optimal number of clusters. The core of this approach is to assess the within cluster sum of squared distance (WCSS) regarding a range of k values (in our case we tested from k=2 to k=10) and identify the “elbow”, corresponding to the optimal number of clusters for the dataset[4,5]. In other words, the “elbow” represents the point where adding more clusters does not significantly improve performance, thus yielding to a balance between model complexity and accuracy. Additionally, we also computed the silhouette score[6] for the same set of k values to observe when the metric significantly decreased compared to the previous value of k[5]. The “elbow” method suggested a best k number of clusters between 4 and 7 and silhouette score method to k=7 (Figure S2). Manual refinements were performed on cluster centroids 1, 2 and 4 to better fit visible cluster boundaries on the PCA plot. We used parameter k = 6 and added manually the 7$^{th}$ cluster, between North-African and European reference clusters, because

Kmeans clustering could not capture this specific subpopulation. We included overlapping SNPs between OFSEP-HD and reference datasets and removed SNPs with high frequency differences between datasets. We also removed AT and GC variants, resulting in 9,326 SNPs used for PCA (Figure S1).

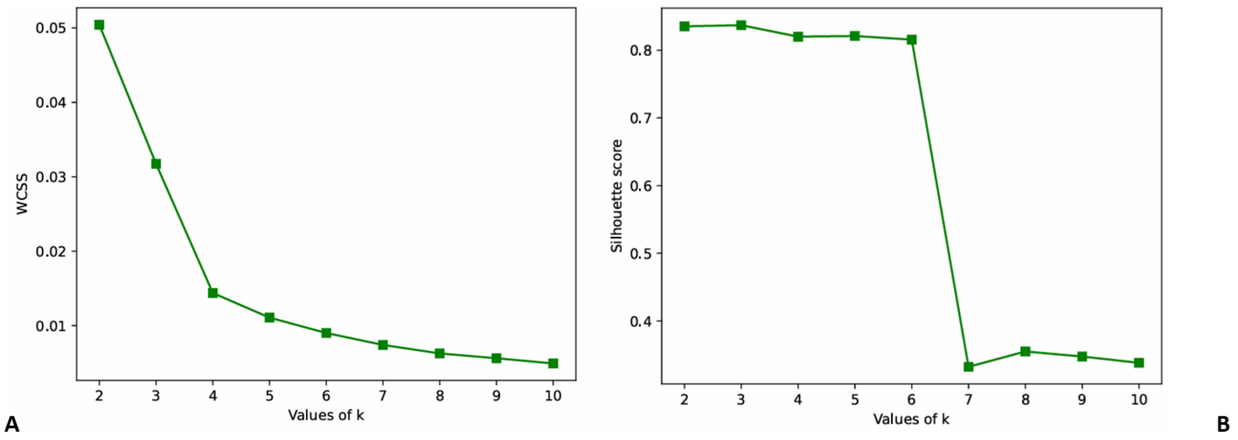


***Figure S2: A:*** *Elbow method plot: within cluster sum of squared distance (WCSS) evolving according to the number of clusters k.* ***B:*** *Silhouette score method plot: silhouette score evolving according to the number of clusters k.*

We used "scikit-learn" and "matplotlib" python libraries to perform K-Means clustering and produce ancestry plots.

**Details on reference population selection and admixture proportions of European, African and East-Asian genome**

During admixture analysis, we selected the most discretizing superpopulation as reference populations, e.g., European, African and East-Asian. The other populations included more recent admixture history. For example, American population from 1000 Genomes show diverse admixture profile with European, African and native American ancestries We used python library "matplotlib" to produce plots and statistics were computed using pandas and NumPy python libraries.

#### Self-reported origins and genetic ancestry: details on comparative statistical analysis

We combined South and East-Asian estimated ancestry clusters into a single Asian cluster for direct comparison with self-reported Asian origins and combined European ancestry cluster with admixed European / North-African ancestry cluster for direct comparison with self-estimated European and France metropole origins (Table S2). We considered this intermediate cluster as an extension of the European reference cluster since the average African genome proportion among this cluster is approximately 11% with a maximum around 14%, lower than average African genome proportion (23.9%) among North-African cluster (Tables S3 – S4 and Figures S3 – S4). Thus, we stay as conservative as possible in the analyses, and avoid including noise in North-African population.

| **Self-reported origins** | **Ancestral cluster** |
|---|---|
| France metropole, Europe | European (clusters 1 + 7) |
| French overseas territories | Afro-Caribbean (cluster 2) |
| North-African | North-African (cluster 3) |
| Sub-Saharan African | African (cluster 5) |
| Asian | East and South-Asian (clusters 4 + 6) |
| Unknown / others | / |

***Table S2****: Mapping between self-reported origin groups and defined ancestry clusters.*

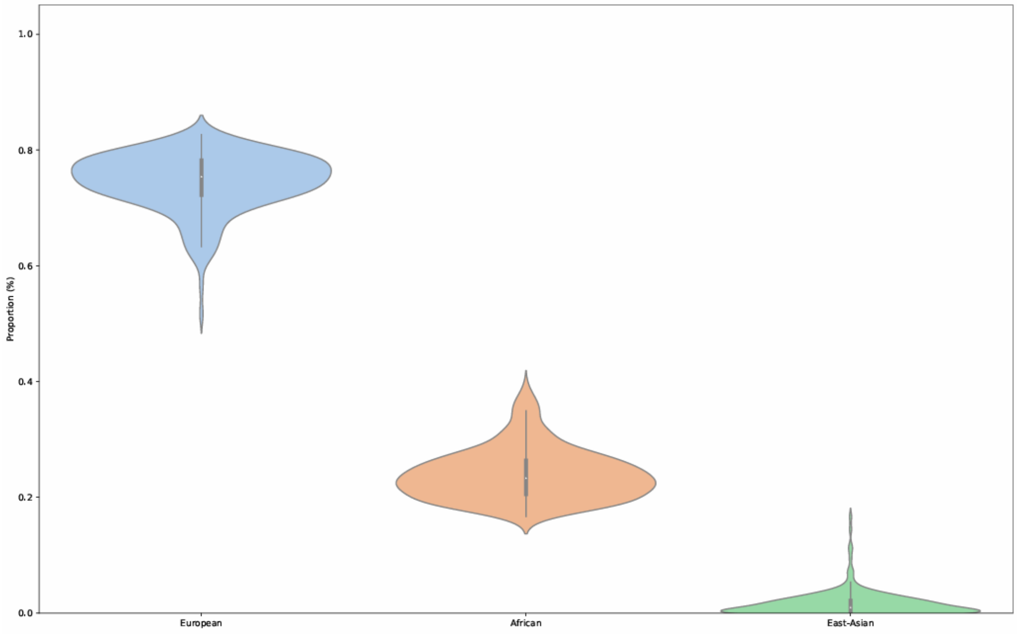


***Figure S3:*** *Violin plot showing European, African and East-Asian genome proportion distributions of the 232 patients from North-African genetic ancestry (cluster 3).*

| Reference population | **Mean** | **Min** | **Max** | **Q1** | **Q3** | **SD** | **N > 90%** | **N > 80%** | **N > 75%** | **N > 50%** | **N > 25%** | **N > 10%** | **N > 5%** |
|---|---|---|---|---|---|---|---|---|---|---|---|---|---|
| **European** | 74.6 % | 51.7 % | 82.7 % | 72.2 % | 78.2 % | 4.9 % | 0 | 21 | 121 | 232 | 232 | 232 | 232 |
| **African** | 23.9 % | 16.7 % | 38.9 % | 20.6 % | 26.3 % | 4.4 % | 0 | 0 | 0 | 79 | 232 | 232 | 232 |
| **East-Asian** | 1.5 % | < 0.01 % | 16.6 % | < 0.01 % | 2.2 % | 2.2 % | 0 | 0 | 0 | 0 | 0 | 4 | 10 |

***Table S3:*** *European, African and East-Asian genome statistic proportion distributions of the 232 patients identified as from North-African ancestry (cluster 3). Q1: 25th percentile; Q3: 75th percentile; SD: Standard Deviation; N: number of patients.*

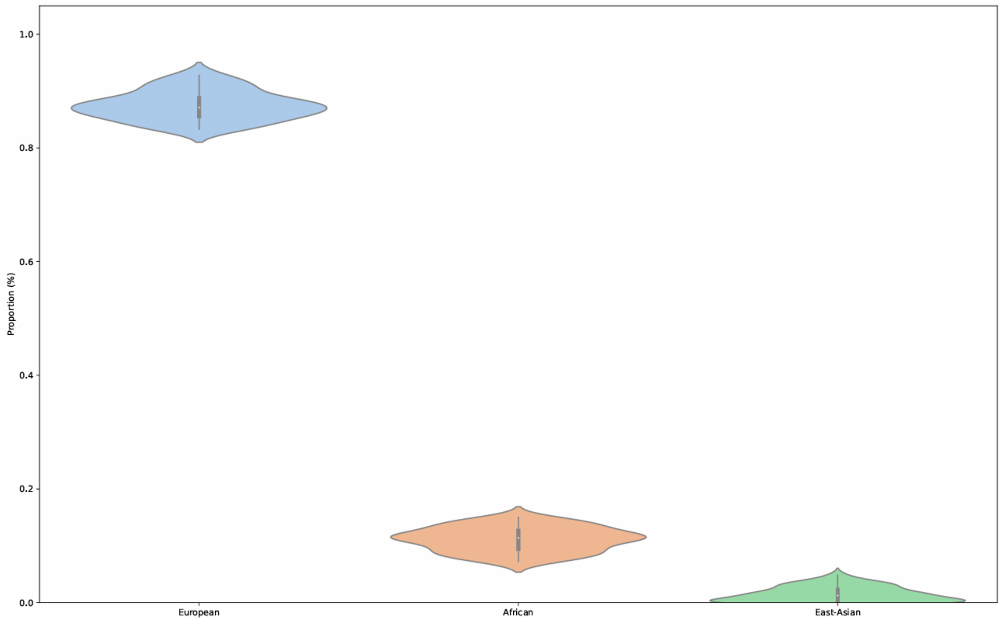


***Figure S4:*** *Violin plot showing European, African and East-Asian genome proportion distributions of the 61 patients from Admixed European / North-African genetic cluster (cluster 7).*

| Reference population | **Mean** | **Min** | **Max** | **Q1** | **Q3** | **SD** | **N > 90%** | **N > 80%** | **N > 75%** | **N > 50%** | **N > 25%** | **N > 10%** | **N > 5%** |
|---|---|---|---|---|---|---|---|---|---|---|---|---|---|
| **European** | 84.7 % | 83.3 % | 92.8 % | 85.6 % | 88.7 % | 2.5 % | 12 | 61 | 61 | 61 | 61 | 61 | 61 |
| **African** | 11.1 % | 7.2 % | 15.0 % | 9.4 % | 12.6 % | 2.1 % | 0 | 0 | 0 | 0 | 0 | 42 | 61 |
| **East-Asian** | 1.4 % | < 0.01 % | 4.9 % | 0.06 % | 2.2 % | 1.3 % | 0 | 0 | 0 | 0 | 0 | 0 | 0 |

***Table S4:*** *European, African and East-Asian genome statistic proportion distributions of the 61 patients from Admixed European / North-African genetic cluster (cluster 7). Q1: $25^{th}$ percentile; Q3: $75^{th}$ percentile; SD: Standard Deviation; N: number of patients.*

We performed whether $X^2$ of independence test or Fisher 's exact test depending on estimated sample size under null hypothesis within each cell of contingency tables, following recommendations from Kim HY et al[7]. The study suggests using Fisher's exact test when more than 20% of cells show small estimated sample size (< 5), and $X^2$ test otherwise. In the case where less than 20% of cells have an expected sample size smaller than 5, we chose to use $X^2$

approximation using Yate's continuity correction[8]. We also computed Cohen's kappa metric, with kappa value of -1 indicating no agreement and 1 suggesting perfect agreement[9,10].

Concerning quantitative statistical analyses of reference genome proportions, we performed several normality tests each tested genome proportion distribution, such as Kolmogorov-Smirnov one-sample test, Anderson-Darling test and even D'Agostino's K-squared test to assess for any significant deviation from normally distributed values with a strong statistical power. When sample size was too small, e.g smaller than 50, we preferred using Shapiro-Wilk test[11]. Non-parametric Mann-Whitney U tests were employed to compare distributions between independent groups where data were not normally distributed in at least one of the compared groups. In the case where both compared distributions followed normal distribution, we preferred using parametric Student t-test making assumption of normal data distribution.

Additionally, we visually compared self-reported origins and genetic ancestry by plotting in black self-reported patients associated to each defined ancestry cluster on the PCA. Tests and figures were implemented using "scikit-learn", "SciPy", and "statsmodels" as well as "NumPy", "matplotlib" and "seaborn" python libraries.

**Details on Polygenic Risk Scores computation**

Log-additive model score used in this study is computed as $\sum_{i=1}^{n}(\text{Asociated Allele Count}(\text{SNP}_\text{i}) \times \ln(\text{OR}_\text{i}))$ with $\text{Associated Allele Count}(\text{SNP}_\text{i})$ the number of OR associated allele for the $i^{th}$ SNP and $\text{OR}_\text{i}$ the Odd-Ratio associated to $\text{SNP}_\text{i}$. Considering the bi-allelic nature of MS associated SNPs, we chose to compute the scores using only risk allele counts and ORs (OR > 1). However, we used HLA alleles ORs whether alleles were associated to protective (OR < 1) or risk effects (OR > 1). SRAC score was defined as a non-weighted by effect size version of the previous model: $\sum_{i=1}^{n}(\text{Risk Allele Count}(\text{SNP}_i))$. All plots and figures were created using python “matplotlib” and “seaborn“ libraries.

| Rsid | HG38 “chr_pos,alt,ref” |
|---|---|
| rs10063294 | 5_35877403,G,A |
| rs1014486 | 3_159973324,C,T |
| rs10191360 | 2_136127109,C,T |
| rs10230723 | 7_50200284,A,T |
| rs10245867 | 7_28102567,T,G |
| rs1026916 | 17_42377817,A,G |
| rs10271373 | 7_139045049,C,A |
| rs1076928 | 6_36380912,T,C |
| rs10801908 | 1_116547871,T,C |
| rs1087056 | 10_31106832,A,G |
| rs10936602 | 3_169818849,C,T |
| rs10951042 | 7_3099783,C,T |
| rs10951154 | 7_27095695,C,T |
| rs11079784 | 17_47624914,T,C |
| rs11083862 | 19_47135282,T,A |
| rs11125803 | 2_24829308,C,T |
| rs1112718 | 10_92719350,G,A |
| rs111430408 | 3_101129753,T,C |
| rs11161550 | 1_85216337,A,G |
| rs111635774 | 6_14690984,T,C |
| rs11231749 | 11_64327706,C,T |
| rs112344141 | 1_155010560,G,T |
| rs11256593 | 10_6075359,C,T |
| **rs114071505** | **6_30092854,G,C** |
| rs11542663 | 6_118894238,C,A |
| rs11578655 | 1_100947346,G,T |
| rs116877451 | 7_50288743,G,A |
| rs116899835 | 14_88057144,T,C |
| rs11749040 | 5_40396323,A,G |
| **rs11751659** | **6_33113855,G,A** |
| rs1177228 | 2_61015275,A,G |
| rs11809700 | 1_92687078,T,C |
| rs11852059 | 14_51839373,C,A |
| rs11899404 | 2_12467767,T,C |
| rs11919880 | 3_32920559,G,A |
| rs12133753 | 1_91756532,T,C |
| rs12147246 | 14_102799507,A,G |
| rs12211604 | 6_7099796,G,A |
| rs12365699 | 11_118872577,A,G |
| rs12434551 | 14_68786647,A,T |
| rs12478539 | 2_43128185,G,C |
| rs1250551 | 10_79299578,T,G |
| rs12588969 | 14_102764421,C,G |
| rs12609500 | 19_11063252,T,C |
| rs12614091 | 2_203768138,T,A |
| rs12622670 | 2_68419404,C,T |

| | |
|---|---|
| rs12722559 | 10_6028310,A,C |
| rs12925972 | 16_79077400,T,C |
| rs12971909 | 19_4466469,A,G |
| rs13066789 | 3_188269836,C,T |
| rs13136820 | 4_40305547,C,T |
| rs1323292 | 1_192571891,G,A |
| rs13327021 | 3_27741524,T,C |
| rs13385171 | 2_65434709,T,C |
| rs1365120 | 11_36416525,C,T |
| rs137955 | 22_39895803,T,C |
| rs138433213 | 3_112975136,G,T |
| rs1399180 | 10_8056756,T,C |
| rs140522 | 22_50532837,T,C |
| rs146566517 | 16_11260022,T,C |
| rs1465697 | 19_49333989,T,C |
| rs149114341 | 11_118912715,A,G |
| rs17051321 | 4_121198294,T,C |
| rs1738074 | 6_159044945,T,C |
| **rs17493811** | **6_32177622,C,G** |
| rs17724508 | 16_79316307,C,T |
| rs17741873 | 10_73894042,T,G |
| rs17780048 | 6_137858009,T,C |
| rs1800693 | 12_6330843,C,T |
| rs1801133 | 1_11796321,A,G |
| rs198398 | 1_11823285,T,C |
| rs2084007 | 5_134555592,T,C |
| rs2150879 | 17_59781849,A,G |
| **rs2229092** | **6_31572980,C,A** |
| rs2248137 | 20_54173204,G,C |
| rs2269434 | 11_47338861,C,T |
| rs2286974 | 16_11020655,G,A |
| rs2289746 | 3_105737111,T,C |
| rs2317231 | 1_157716547,T,G |
| rs2327586 | 6_135174088,T,C |
| rs2331964 | 3_121824051,T,C |
| rs2364485 | 12_6405797,A,C |
| rs244656 | 5_134114136,A,T |
| rs2469434 | 18_69876810,C,T |
| rs249677 | 5_142159774,C,A |
| **rs2523500** | **6_31550577,G,A** |
| rs2546890 | 5_159332892,G,A |
| rs2585447 | 20_54127898,C,T |
| rs2590438 | 3_187848180,G,T |
| rs2705616 | 4_86941244,C,G |
| rs2726479 | 4_105334432,T,C |
| rs2836438 | 21_38492803,A,G |
| **rs2844482** | **6_31571990,T,C** |
| rs28703878 | 8_78504987,G,A |
| rs28834106 | 19_10481468,C,T |
| rs2986736 | 1_6452487,C,T |
| **rs3093982** | **6_31529467,A,G** |
| **rs3097671** | **6_33079835,C,G** |

| | |
|---|---|
| **rs3135024** | **6_33079689,C,T** |
| rs3184504 | 12_111446804,C,T |
| rs32658 | 5_119367967,T,G |
| rs34026809 | 11_118609980,G,C |
| rs34536443 | 19_10352442,G,C |
| rs34681760 | 5_6712721,T,C |
| rs34695601 | 14_75547955,C,T |
| rs34723276 | 1_100824876,G,A |
| rs34947566 | 16_11319069,A,C |
| rs354033 | 7_149592373,A,G |
| rs35486093 | 1_85264137,G,A |
| rs35540610 | 2_230257114,C,T |
| rs35703946 | 16_85987899,A,G |
| rs3737798 | 1_160420194,G,A |
| rs3809627 | 16_30091839,A,C |
| rs3923387 | 8_143912625,T,C |
| rs405343 | 16_1017832,T,G |
| rs4262739 | 11_128551280,A,G |
| rs4325907 | 3_102030178,C,T |
| rs438613 | 3_28030595,T,C |
| rs4409785 | 11_95578258,C,T |
| rs4728142 | 7_128933913,A,G |
| rs4796224 | 17_36486677,A,G |
| rs4808760 | 19_18191169,G,C |
| rs4812772 | 20_43950411,T,C |
| rs483180 | 1_119724882,C,G |
| rs4896153 | 6_135512325,A,T |
| rs4939490 | 11_61026179,G,C |
| rs4940730 | 18_58602505,G,A |
| rs531612 | 11_65937961,C,T |
| rs56095240 | 11_95688666,T,A |
| rs570429157 | 11_14846770,G,A |
| rs57116599 | 2_112013222,A,G |
| rs5756405 | 22_36914912,G,A |
| rs58166386 | 19_16448610,G,A |
| rs58394161 | 1_92474402,C,T |
| rs59655222 | 1_200906769,C,T |
| rs6020055 | 20_49805558,G,A |
| rs6032662 | 20_46105671,C,T |
| rs60600003 | 7_37342861,G,T |
| rs6072343 | 20_41339548,A,G |
| rs61708525 | 12_94267677,G,A |
| rs61863928 | 10_62689789,T,G |
| rs61884005 | 11_14381384,C,G |
| rs62013236 | 15_78955140,T,C |
| rs62420820 | 6_137116920,A,G |
| rs631204 | 6_137638318,A,C |
| rs6427540 | 1_160664798,T,C |
| rs6496663 | 15_90344352,C,A |
| rs6533052 | 4_102990624,G,A |
| rs6564681 | 16_79618823,C,T |
| rs6589706 | 11_118877104,A,G |

| | |
|---|---|
| rs6589939 | 11_122647817,G,A |
| rs6670198 | 1_2589088,C,T |
| rs6672420 | 1_24964519,A,T |
| rs67111717 | 5_177363161,G,A |
| rs6738544 | 2_191124630,A,C |
| rs6742 | 20_63743088,T,C |
| rs6789653 | 3_141432148,A,G |
| rs67934705 | 1_23881014,A,G |
| rs6837324 | 4_48125245,G,A |
| rs6880809 | 5_40429148,A,T |
| rs6911131 | 6_143544084,G,A |
| rs6990534 | 8_127801845,A,G |
| rs701006 | 12_57713053,A,G |
| rs719316 | 6_16672529,C,T |
| rs7222450 | 17_45330304,A,G |
| rs7260482 | 19_44640642,C,A |
| rs72922276 | 1_64963636,A,G |
| rs72928038 | 6_90267049,A,G |
| rs72989863 | 4_163572655,A,G |
| rs73414214 | 7_106066016,A,C |
| rs735542 | 8_127163451,G,A |
| **rs7454108** | **6_32713706,C,T** |
| rs75191738 | 6_130027112,T,C |
| rs75937181 | 3_122064168,G,T |
| rs760517 | 22_36862944,T,C |
| rs7731626 | 5_56148856,A,G |
| rs77654077 | 13_99374698,C,A |
| rs7819665 | 8_128165523,C,T |
| rs7855251 | 9_98105907,C,T |
| rs78727559 | 8_94839590,G,T |
| rs7975763 | 12_123119506,T,C |
| rs7977720 | 12_9713753,T,C |
| rs79979643 | 1_32272814,A,G |
| rs802730 | 6_127958959,C,T |
| rs8062446 | 16_57043182,T,C |
| rs883871 | 17_40096407,A,G |
| **rs9266629** | **6_31379045,C,T** |
| **rs9271366** | **6_32619077,G,A** |
| **rs9277626** | **6_33114046,G,A** |
| rs9308424 | 1_212704434,A,G |
| rs9568402 | 13_50387821,A,T |
| rs9591325 | 13_50237084,C,T |
| rs9610458 | 22_21851064,C,T |
| rs962052 | 2_150787689,C,T |
| rs9808753 | 21_33415005,G,A |
| rs983494 | 1_160734175,A,G |
| rs9843355 | 3_119509661,A,G |
| rs9863496 | 3_18757356,C,T |
| rs9878602 | 3_71486187,G,T |
| rs9900529 | 17_75339695,C,G |
| rs9909593 | 17_39813896,G,A |
| rs9955954 | 18_58680812,G,A |

| | |
|---|---|
| rs9992763 | 4_108137562,G,T |
| **HLA alleles** | |
| **HLA-DRB1*15:01** | |
| **HLA-B*38:01** | |
| **HLA-DRB1*13:03** | |
| **HLA-DRB1*01:03** | |
| **HLA-B*52:01** | |
| **HLA-B*35:03** | |
| **HLA-DRB1*04:11** | |

***Table S5:*** *210 MS (190 non-MHC autosomal SNPs + 13 MHC SNPs + 7 HLA alleles) associated susceptibility variants available out of the 8.5 million variants of post-imputed and post-processed datasets. Note 233 MS susceptibility variants are known to date[12]. Bold variants represent MHC region variants.*

### Details on synthetic data generation

Three main GDPR criterias to consider a dataset as anonymous:

None of those criteria should be possible in an anonymous dataset. We also described below the different privacy metrics associated to each criterion.

- **Singling-out**: risk to identify a patient in the dataset.
  - ➔ **Hidden Rate (HR):** probability that an attacker makes a mistake when trying to link a patient with its most similar synthetic patient.
  - ➔ **Median Local Cloaking (LC):** median number of synthetic patients closer to a given original patient than its own associated synthetic patient it was generated from.
- **Linkability**: capacity to link patients with another dataset.
  - ➔ **Median Distance to Closest (DTC):** median distance between an original patient and the closest synthetic patient in the reduced space.
  - ➔ **Distance to Closest Ratio (DCR):** distance between synthetic record and its closest original record divided by the distance to its second closest original record.
  - ➔ **Column direct match protection:** how unlikely each column in the data might be used as a direct identifier.

- ➔ **Row direct match protection:** proportion of patients in original data that we can bound identical in synthetic data.

- **Inference:** probabilistic deduction of some patient features values.

The “Avarization”[13] methodology used to generate the synthetic OFSEP-HD genetic dataset was initially developed by Octopize, a French, Nantes’ startup company (https://www.octopize.io). The method allows patient centric synthetic data generation based on a local model established from K-Nearest Neighbors algorithm (KNN) of a given patient to create anonymized patients while keeping the initial distribution statistics as well as the informativeness of the original dataset, thus strongly limiting risk of re-identification through genetic data. This methodology is in compliance with European data privacy regulations and was validated by GDPR regulations.

We selected 82 features, such as quality control features, principal components coordinate, ancestry clusters, PRS score values, proportions of African, European and Asian genome, HLA alleles and SNPs. Pre-avatarization processing was designed to remove variables computed from other variables and create the categorical encoding of HLA-alleles and SNPs. Out of a set of avatarization parameters combinations, we first of all selected datasets from all combinations that yielded sufficient privacy metrics (custom thresholds: LC >= 3, HR ≈ 85%). Out of all remaining combinations, we selected the one yielding the best fidelity to original data. For each avatar dataset, we assessed the number of categorical variables with at least one missing modality. We computed Hellinger and Gower’s distances and performed permutation test on Gower’s distance. Moreover, we performed univariate statistical analysis of each avatar dataset compared to the original one and corrected p-values for multiple testing using Bonferroni correction. After defining the best avatar model in a single generation configuration, we averaged the generations from multiple avatar models using the best previously defined configuration but from various random seeds. A rule-based post-generation processing was implemented to compute back calculated features (East Asian genome proportions) and to ensure respect of biological and genetic constraints in the generated dataset. For example, when a synthetic patient had more than 2 alleles associated to a specific HLA locus, we chose to firstly remove most frequent alleles to limit the number of alleles to 2 for the given locus of the synthetic patient. The aim of this methodology is to balance the underestimation of rare modalities caused by the avatarization method[13]. Although the entire OFSEP-HD population was compiled using this method, we selected a subset of 1,000 patients to be included in the final open shared dataset.

**Ancestry analysis: Principal Components Analysis of OFSEP-HD MS patients**

| PC | chr_pos,alt,ref | explained_variance (%) | rsid |
|---|---|---|---|
| **1** | 1_234157906,G,A | 0.4977722879225727 | rs6586371 |
| **1** | 11_130139411,A,G | 0.479345690590744 | rs7103088 |
| **1** | 6_94748599,A,G | 0.4785832107011511 | rs794672 |
| **1** | 7_6206892,T,C | 0.4597753734245259 | rs6962026 |
| **1** | 5_142715846,C,T | 0.4553275740685673 | rs12516796 |
| **1** | 17_263243,T,C | 0.4477027751726382 | rs7208188 |
| **1** | 14_63325784,C,T | 0.4468132153014464 | rs1268646 |
| **1** | 15_29024263,A,G | 0.4421112559822902 | rs8031475 |
| **1** | 1_244297524,T,G | 0.4407133761847031 | rs7554059 |
| **1** | 7_115735678,C,T | 0.4382988565343255 | rs6946417 |
| 2 | 8_138027346,G,A | 0.501076367444142 | rs6997563 |
| 2 | 7_113642561,G,A | 0.4670189323756585 | rs1858892 |
| 2 | 2_28270842,A,C | 0.4654939725964728 | rs4666050 |

| | | | |
|---|---|---|---|
| 2 | 13_106183039,A,G | 0.4554546540501661 | rs9519977 |
| 2 | 2_176880003,A,G | 0.4377905366079302 | rs17626597 |
| 2 | 12_49477181,G,A | 0.4321990174175822 | rs11169042 |
| 2 | 4_85727592,A,G | 0.4299115777488035 | rs41527748 |
| 2 | 1_26725589,A,G | 0.4215242989632816 | rs1376925057 |
| 2 | 7_72120720,C,A | 0.4097058606745914 | rs1808315010 |
| 2 | 16_5548925,A,C | 0.4097058606745914 | rs11076930 |
| **3** | 1_51318915,T,C | 0.6205315501470317 | rs11577254 |
| **3** | 8_80669869,C,T | 0.5627101585195691 | rs12547769 |
| **3** | 13_27373838,C,T | 0.4939598884746082 | rs11616513 |
| **3** | 3_157097491,G,A | 0.4924349286954223 | rs13094970 |
| **3** | 10_10618898,G,A | 0.4784561307195523 | rs11256710 |
| **3** | 5_147181853,C,T | 0.4668918523940598 | rs17582364 |
| **3** | 1_110227671,A,G | 0.4516422546022016 | rs2077402 |
| **3** | 11_30669992,C,T | 0.4477027751726382 | rs1447265 |
| **3** | 4_164643363,A,G | 0.439442576368715 | rs6824404 |
| **3** | 3_159996059,C,T | 0.4369009767367386 | rs640039 |
| 4 | 13_50465285,G,A | 0.5860928751337517 | rs1666992257 |
| 4 | 20_38277492,C,T | 0.5352608824942242 | rs2076504439 |
| 4 | 2_176393268,G,A | 0.4994243276833574 | rs16863769 |
| 4 | 11_92986515,T,C | 0.4943411284194047 | rs1858194907 |
| 4 | 2_163612849,C,T | 0.4937057285114105 | rs16848713 |
| 4 | 9_18280145,A,G | 0.4915453688242306 | rs1055151626 |
| 4 | 8_102301752,C,T | 0.4874788094130683 | rs10505024 |
| 4 | 7_34093438,A,G | 0.4776936508299594 | rs973766 |
| 4 | 22_41814981,C,T | 0.4690522120812397 | rs139568 |
| 4 | 14_46457195,T,G | 0.4667647724124609 | rs1761031 |
| **5** | 4_38909584,G,A | 0.7007190185358861 | rs16994919 |

| | | | |
|---|---|---|---|
| **5** | 19_37387954,A,G | 0.5066678866344901 | rs3826849 |
| **5** | 12_128179388,A,G | 0.5003138875545491 | rs11059562 |
| **5** | 22_26192779,T,C | 0.4600295333877235 | rs134765 |
| **5** | 16_8287386,G,A | 0.4571066938109508 | rs17145680 |
| **5** | 14_54939405,G,A | 0.4468132153014464 | rs10140164 |
| **5** | 3_77285851,G,A | 0.4389342564423197 | rs10433496 |
| **5** | 2_135998114,A,G | 0.4348676970311575 | rs1683008843 |
| **5** | 10_75699317,T,C | 0.4235575786688627 | rs11001475 |
| **5** | 6_116946201,C,T | 0.4229221787608684 | rs1741692 |
| 6 | 4_38801442,G,A | 0.7369368132915496 | rs5743592 |
| 6 | 20_59179868,T,C | 0.6420080670372319 | rs11086667 |
| 6 | 11_48150813,T,C | 0.5079386864504782 | rs17198978 |
| 6 | 1_187105358,C,T | 0.4989160077569621 | rs10798086 |
| 6 | 8_118094812,A,G | 0.4938328084930093 | rs11992017 |
| 6 | 5_134082114,A,G | 0.4901474890266436 | rs10900826 |
| 6 | 22_45457685,G,A | 0.4895120891186495 | rs5765377 |
| 6 | 10_79252314,G,A | 0.4766770109771688 | rs11002893 |
| 6 | 8_4191489,T,C | 0.4605378533141188 | rs1437882730 |
| 6 | 22_42218356,C,T | 0.4601566133693224 | rs932376 |
| **7** | 2_135998114,A,G | 1.1718045103227068 | rs1683008843 |
| **7** | 17_35212806,C,T | 0.7078354975054201 | rs1383541 |
| **7** | 2_31231679,T,C | 0.6994482187198979 | rs605832 |
| **7** | 18_6602285,A,C | 0.6839444609648421 | rs4508504 |
| **7** | 20_6001958,C,T | 0.6514119856755446 | rs1191991563 |
| **7** | 2_235666672,G,A | 0.6182441104782529 | rs1806688 |
| **7** | 18_48714340,C,A | 0.6176087105702588 | rs9947954 |
| **7** | 3_14776220,A,G | 0.601977872833604 | rs3899614 |
| **7** | 3_125649018,G,A | 0.6009612329808135 | rs1481387280 |

| | | | |
|---|---|---|---|
| **7** | 7_83328461,T,C | 0.5858387151705541 | rs2371662 |
| 8 | 21_37969257,T,C | 0.5344984026046314 | rs2836072 |
| 8 | 2_118803661,A,G | 0.5266194437455046 | rs12988660 |
| 8 | 20_13162915,C,A | 0.5153093253832097 | rs7264061 |
| 8 | 6_166489859,C,T | 0.5131489656960299 | rs1781530420 |
| 8 | 6_71922044,A,G | 0.4911641288794342 | rs6931045 |
| 8 | 1_179833243,T,C | 0.4868434095050743 | rs373411017 |
| 8 | 16_89442454,C,A | 0.4704500918788267 | rs9302767 |
| 8 | 7_147520914,G,A | 0.4665106124492634 | rs2888493 |
| 8 | 1_210907839,C,T | 0.456217133939759 | rs1697621 |
| 8 | 7_67593785,A,G | 0.4465590553382487 | rs10270452 |
| **9** | 2_21719863,T,C | 0.4915453688242306 | rs11683091 |
| **9** | 6_71278507,G,A | 0.4895120891186495 | rs6907281 |
| **9** | 12_125427265,G,A | 0.4818872902227203 | rs366660 |
| **9** | 3_193412595,A,G | 0.4611732532221129 | rs6764121 |
| **9** | 16_89541006,A,G | 0.4601566133693224 | rs2058487249 |
| **9** | 5_147349399,G,A | 0.4469402952830452 | rs17482628 |
| **9** | 1_84266846,A,G | 0.4399508962951102 | rs12145396 |
| **9** | 13_27515408,A,G | 0.4370280567183374 | rs12872402 |
| **9** | 16_76235062,C,T | 0.4343593771047622 | rs11861749 |
| **9** | 22_48317415,C,T | 0.4285136979512165 | rs133646 |
| 10 | 10_2021507,T,C | 0.5466980808381179 | rs7070363 |
| 10 | 10_1984214,C,T | 0.5455543610037286 | rs1010571 |
| 10 | 14_101497049,G,A | 0.5022200872785314 | rs1889364 |
| 10 | 12_105048408,C,T | 0.4939598884746082 | rs10746009 |
| 10 | 18_38119474,T,C | 0.4931974085850152 | rs4799475 |
| 10 | 9_116249112,A,G | 0.4798540105171393 | rs10983088 |
| 10 | 13_48660330,C,A | 0.4787102906827499 | rs4942769 |

| | | | |
|---|---|---|---|
| 10 | 4_146976521,T,C | 0.4741354113451924 | rs7663340 |
| 10 | 14_40920099,A,G | 0.4666376924308621 | rs1154143 |
| 10 | 2_82561682,C,T | 0.4438903757246736 | rs7581293 |
| **11** | 10_66215237,A,C | 0.5287798034326844 | rs1037988 |
| **11** | 9_18682354,C,A | 0.5014576073889385 | rs2779127 |
| **11** | 14_32002883,T,C | 0.4916724488058295 | rs1013251 |
| **11** | 3_8054139,G,A | 0.4865892495418767 | rs1111569 |
| **11** | 11_46177290,C,T | 0.4837934899467026 | rs11038804 |
| **11** | 9_4228550,C,T | 0.4679084922468503 | rs2890551 |
| **11** | 8_117223802,C,T | 0.4615544931669094 | rs7828267 |
| **11** | 5_112572261,G,A | 0.4571066938109508 | rs9790990 |
| **11** | 2_240670162,C,T | 0.4526588944549921 | rs2043751786 |
| **11** | 5_31589373,C,T | 0.4441445356878712 | rs6891910 |
| 12 | 14_76033960,C,T | 0.5175967650519886 | rs9323624 |
| 12 | 15_51387273,T,C | 0.5108615260272511 | rs12899040 |
| 12 | 14_89657163,C,T | 0.5055241668001007 | rs2181663 |
| 12 | 9_131595766,T,C | 0.4790915306275464 | rs3808803 |
| 12 | 1_208281026,T,C | 0.4766770109771688 | rs1781034 |
| 12 | 18_24628637,A,G | 0.4696876119892338 | rs339866 |
| 12 | 1_40620552,T,C | 0.4676543322836528 | rs12125724 |
| 12 | 14_76117432,C,T | 0.4474486152094405 | rs2591078 |
| 12 | 6_129716775,T,C | 0.445034095559063 | rs12205840 |
| 12 | 4_168639514,G,A | 0.4416029360558949 | rs6552978 |
| **13** | 9_7007063,A,G | 0.5211550045367555 | rs7025832 |
| **13** | 15_47664559,A,G | 0.5080657664320771 | rs1223793888 |
| **13** | 9_101788272,G,A | 0.4882412893026613 | rs1885387 |
| **13** | 11_115220399,G,T | 0.4812518903147262 | rs4936323 |
| **13** | 6_155434840,G,A | 0.4788373706643487 | rs231959 |

| | | | |
|---|---|---|---|
| **13** | 2_219829393,G,T | 0.4757874511059771 | rs17609914 |
| **13** | 20_44407812,A,G | 0.4755332911427794 | rs8114057 |
| **13** | 14_98257481,T,C | 0.4705771718604255 | rs7149283 |
| **13** | 9_4237546,G,A | 0.4689251320996409 | rs2224492 |
| **13** | 8_5923263,T,G | 0.4675272523020539 | rs1798961463 |
| 14 | 7_24309394,C,T | 0.5254757239111152 | rs1298761523 |
| 14 | 20_19245440,A,G | 0.5141656055488204 | rs7265772 |
| 14 | 20_19240267,A,C | 0.5084470063768736 | rs3790167 |
| 14 | 3_122716903,A,G | 0.5055241668001007 | rs2063515 |
| 14 | 2_58466715,A,G | 0.496120248161788 | rs6742732 |
| 14 | 11_4478801,G,A | 0.4766770109771688 | rs12796582 |
| 14 | 9_111624855,G,A | 0.4524047344917944 | rs7875749 |
| 14 | 14_38238912,A,G | 0.440077976276709 | rs5001880 |
| 14 | 9_78892320,A,G | 0.4391884164055173 | rs11137945 |
| 14 | 9_81595294,T,C | 0.4357572569023492 | rs7860431 |
| **15** | 10_66040917,T,C | 0.7129186967693728 | rs1903867 |
| **15** | 18_3176065,T,C | 0.6218023499630198 | rs2230162 |
| **15** | 5_56347016,C,T | 0.5724953171026781 | rs10052062 |
| **15** | 18_3210636,C,T | 0.5527979199548612 | rs8090956 |
| **15** | 12_118610142,G,A | 0.5436481612797462 | rs2730564 |
| **15** | 10_66211296,G,A | 0.5173426050887908 | rs10822750 |
| **15** | 14_65258452,G,A | 0.5088282463216699 | rs8003310 |
| **15** | 5_45734981,T,C | 0.5071762065608854 | rs6451814 |
| **15** | 10_25634336,T,C | 0.5026013272233278 | rs4237376 |
| **15** | 18_3224542,A,G | 0.4987889277753632 | rs7238320 |
| 16 | 4_69613253,C,T | 0.510226126119257 | rs6848997 |
| 16 | 7_23547026,T,C | 0.5065408066528913 | rs13236851 |
| 16 | 14_85616935,C,T | 0.4836664099651038 | rs1950956 |

| | | | |
|---|---|---|---|
| 16 | 10_10372231,A,C | 0.4780748907747558 | rs1302535298 |
| 16 | 2_29312859,A,G | 0.4663835324676645 | rs6708344 |
| 16 | 14_85724245,T,C | 0.4619357331117058 | rs7140553 |
| 16 | 12_118610142,G,A | 0.4545650941789743 | rs2730564 |
| 16 | 7_24226485,G,A | 0.4521505745285968 | rs10268827 |
| 16 | 10_66040917,T,C | 0.4489735749886263 | rs1903867 |
| 16 | 6_97711860,C,T | 0.4483381750806323 | rs1328797778 |
| **17** | 12_52205638,T,C | 0.565505918114743 | rs12369653 |
| **17** | 13_47558599,C,T | 0.5353879624758231 | rs12877713 |
| **17** | 18_60249319,C,A | 0.5109886060088499 | rs35623600 |
| **17** | 13_47558353,C,A | 0.4933244885666141 | rs9534681 |
| **17** | 16_56447370,A,G | 0.4651127326516764 | rs4784669 |
| **17** | 2_54718637,G,A | 0.4619357331117058 | rs2248749 |
| **17** | 3_120284297,A,G | 0.4592670534981307 | rs9843220 |
| **17** | 12_118601920,A,G | 0.4585045736085378 | rs1520780 |
| **17** | 5_40790449,A,G | 0.4573608537741484 | rs10074991 |
| **17** | 7_38385171,T,C | 0.4499902148414169 | rs1195181915 |
| 18 | 18_24614444,A,G | 0.5590248390532033 | rs746622 |
| 18 | 4_17476645,A,G | 0.5498750803780884 | rs4698599 |
| 18 | 11_55728146,C,T | 0.5333546827702419 | rs295595 |
| 18 | 11_56566981,A,G | 0.5327192828622479 | rs11607910 |
| 18 | 18_24628637,A,G | 0.5084470063768736 | rs339866 |
| 18 | 2_58605272,G,A | 0.4925620086770212 | rs17269126 |
| 18 | 22_27724719,T,C | 0.4761686910507735 | rs7285204 |
| 18 | 5_81626930,G,A | 0.4685438921548444 | rs7707981 |
| 18 | 5_30999623,T,C | 0.4653668926148739 | rs12519137 |
| 18 | 15_86297613,G,A | 0.4591399735165318 | rs33987779 |
| **19** | 15_86297613,G,A | 0.7861167661702924 | rs33987779 |

| | | | |
|---|---|---|---|
| **19** | 15_86145592,G,A | 0.5358962824022183 | rs16948699 |
| **19** | 15_87497169,C,T | 0.5203925246471625 | rs12594783 |
| **19** | 15_86188466,G,A | 0.4962473281433868 | rs8027839 |
| **19** | 7_39051098,C,A | 0.4884954492658589 | rs12531458 |
| **19** | 6_24466493,A,G | 0.4818872902227203 | rs2244216 |
| **19** | 4_87870427,T,C | 0.4756603711243782 | rs8180170 |
| **19** | 2_3674904,G,A | 0.4628252929828976 | rs13409287 |
| **19** | 6_74555280,C,T | 0.4578691737005437 | rs1577217 |
| **19** | 3_13298510,A,G | 0.4474486152094405 | rs357121 |
| 20 | 14_98257481,T,C | 0.6714906227681579 | rs7149283 |
| 20 | 13_78441535,T,C | 0.6464558663931905 | rs1881211542 |
| 20 | 14_98263417,C,A | 0.5805013559434037 | rs10141969 |
| 20 | 18_37906950,C,T | 0.506032486726496 | rs4611540 |
| 20 | 3_159275450,C,T | 0.4829039300755109 | rs1448996 |
| 20 | 5_133669091,C,T | 0.4803623304435346 | rs1860207 |
| 20 | 17_27766289,C,T | 0.4787102906827499 | rs2297514 |
| 20 | 15_54241689,A,G | 0.4701959319156291 | rs12912762 |
| 20 | 11_94229493,C,T | 0.4577420937189448 | rs2399822 |
| 20 | 13_78442097,G,A | 0.4550734141053696 | rs9593324 |

***Table S6:*** *Ten most impacting SNP for each of the 20 principal components from ancestry analysis. chr: Chromosome in hg38 genome build; pos: position in hg38 genome build; alt: alternative allele in OFSEP-HD dataset; ref: reference allele in OFSEP-HD dataset. Alternative and reference alleles may be switched depending on considered reference database.*

### Other demographic and clinical description of OFSEP-HD cohort

Patients presented a mean of 42.34 years old (Table S7). The average age of onset is around 31.18 years old. The Expanded Disability Status Scale (EDSS) measures the severity of disability considering several modalities (walk, cognitive tests, sight …) on a non-linear scale with 0.5 steps starting from 0 (no disability) to 10 (death). EDSS is in average in between 2 and 3 at inclusion. Relapsing-Remitting form of the disease is the main phenotype in the cohort (Table S8). Moreover, sex distribution among the cohort reflects the global distribution of the disease across sex, with around 73.72 % of women and 26.28 % of men (Table S9)[14,15]. Python 3 with pandas and numpy libraries were employed for the demographic description of the OFSEP-HD cohort.

| | **Mean** | **Min** | **Max** | **Q1** | **Q3** | **SD** |
|---|---|---|---|---|---|---|
| **Age inclusion** | 42.34 | 16 | 83 | 34 | 51 | 11.53 |
| **Age onset** | 31.18 | 4 | 73 | 24 | 38 | 10.10 |
| **EDSS inclusion** | 2.40 | 0 | 7 | 1 | 4 | 1.94 |

***Table S7****: Age at onset and disability at inclusion of 2542 remaining OFSEP-HD patients after quality controls and processing steps.*

| | **Count** | **%** |
|---|---|---|
| **CIS** | 379 | 14.91 |
| **PP** | 149 | 5.86 |
| **RR** | 1,711 | 67.31 |
| **SP** | 302 | 11.88 |
| **Unknown** | 1 | 0.04 |

***Table S8:*** *Phenotype at inclusion of 2542 remaining OFSEP-HD patients after quality controls and processing steps.*

| | Count | % |
|---|---|---|
| **Female** | 1,874 | 73.72 |
| **Male** | 668 | 26.28 |

***Table S9:*** *Self-reported sex of 2,542 remaining OFSEP-HD patients after quality controls and processing steps.*

### Admixture proportions of European, African and East-Asian genome in OFSEP-HD cohort

| Reference population | **Mean** | **Min** | **Max** | **Q1** | **Q3** | **N > 85 %** | **N > 80%** | **N > 75%** | **N > 50%** | **N > 25%** | **N > 10%** | **N > 5%** |
|---|---|---|---|---|---|---|---|---|---|---|---|---|
| **European** | 93.7 % | < 0.01 % | 99.9 % | 95.3 % | 98.7 % | 2,231 | 2,274 | 2,379 | 2,506 | 2,522 | 2,533 | 2,536 |
| **African** | 4.6 % | < 0.01 % | 99.9 % | 0.01 % | 2.8 % | 13 | 14 | 16 | 27 | 115 | 314 | 382 |
| **East-Asian** | 1.7 % | < 0.01 % | 99.3 % | < 0.01 % | 2.3 % | 2 | 2 | 2 | 4 | 10 | 42 | 101 |

***Table S10:*** *European, African and East-Asian genome proportion distributions statistics of the global population of 2542 OFSEP-HD patients. N: number of patients.*

### An underestimation of North-African self-reported origins compared to North-African genetic ancestry

Findings indicate a significant and strong relationship between self-reported North-African origins and North-African genetic ancestry ($X^2$ without Yate's correction, $p < 0.0001$), meaning that agreement is not due to chance, enabling comparisons (Table S11).

| | **Self-reported North-African origins** | **Other self-reported origins** | **Total** |
|---|---|---|---|
| **North-African genetic ancetry** | 112 | 120 | 232 |
| **Other genetic ancestry** | 15 | 2,295 | 2,310 |
| **Total** | 127 | 2,415 | 2,542 |

***Table S11****: Contingency table of North-African self-reported individual origins' status compared to patients North-African genetic ancestry status. All expected frequencies are > 5 in all cells of the expected frequencies table (data not shown) meaning* *that we can use a $X^2$ test without Yate's correction to compare patients' North-African origin self-reporting and genetic ancestry status.*

The assessed agreement is further supported by the fact that North-African self-reported patients show important sensitivity of 88.19%, with 112 patients from North-African genetic ancestry out of the 127 patients who self-reported North-African origins. This is complemented by a high specificity of 99.35%, with the large majority of patients from other genetic ancestry (2295/2310) self-reporting other origins. Those results indicate that patients who self-reported other origins are usually marked as from other genetic ancestry cluster. Fifteen patients did not self-report North-African origins while they were genetically estimated from North-African ancestry. Thirteen of those patients are genetically classified in the European cluster (cluster 1 and 7) and two of them are clustered in African cluster (cluster 5) (Figure S6). This may indicate a tendency that discordant North-African self-reporting and genetic ancestry is more frequent for patients with a greater European genomic contribution than for patients with a more pronounced African genome proportion. Those results further highlight the genetic proximity between North-African and European ancestries.

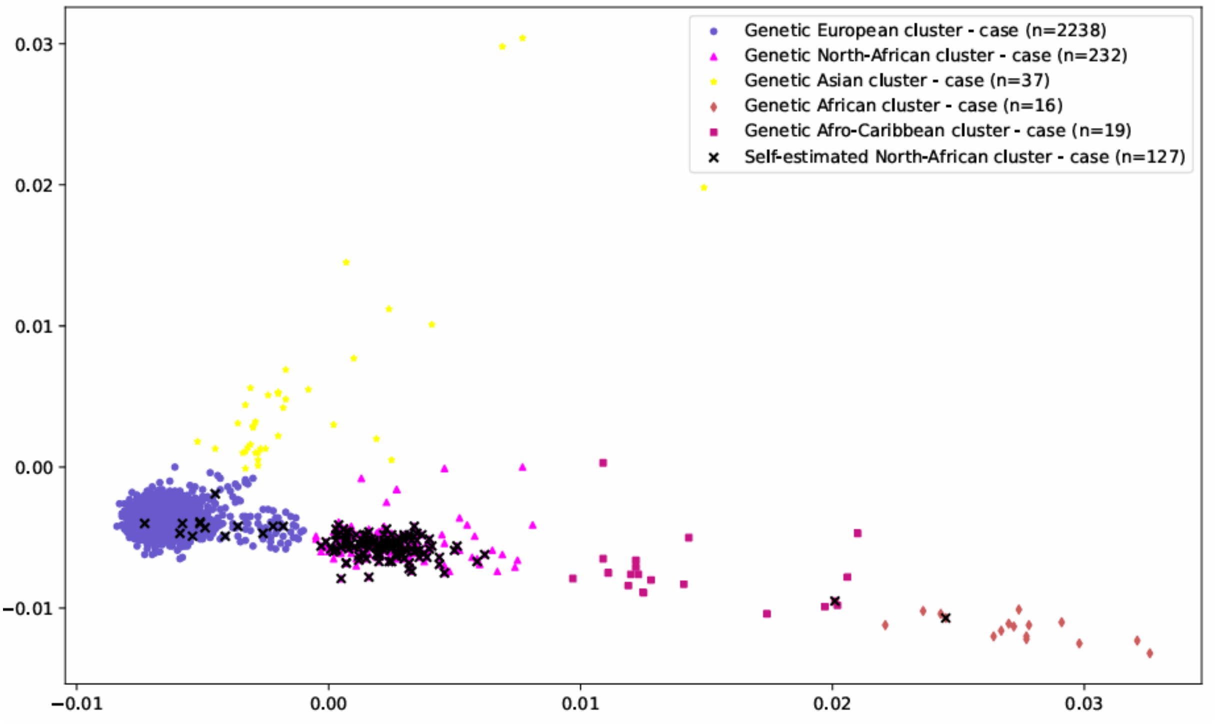


***Figure S5:*** *Self-reported North-African origins distribution (represented as black stars) displayed on the two first components of the PCA from the ancestry analysis. Yellow: Asian cluster, blue-violet: European cluster, pink: North-African cluster, fuchsia: Afro-Caribbean cluster, African cluster.*

Proportion of discordant pairs: (120 + 15) / 2542 * 100 = 5.31 %

### Other self-reported origins and genetic ancestries

#### *African ancestry*

African gen²etic ancestry (cluster 5) was compared to Sub-Saharan-African self-reported origins. This cluster represent the group with the strongest African ancestry.

| | Sub-Saharan African self-reported origins | Other self-reported origins | Total |
|---|---|---|---|
| **African genetic ancestry** | 5 | 11 | 16 |
| **Other genetic ancestry** | 5 | 2,521 | 2,526 |
| **Total** | 10 | 2,532 | 2,542 |

***Table S12:*** *Contingency table of Sub-Saharan-African self-reported individual origins' status compared to patients' African (cluster 5) genetic ancestry status. All expected frequencies are not > 5 in all cells of the expected frequencies table (data not shown). One cell (25%) shows very low sample size, indicating that we need to use an exact Fisher test.*

Fisher's exact test resulted in a significant association between Sub-Saharan-African reporting and African genetic ancestry ($p < 0.001$). However, 68.75 % (11/16) of patients from African genetic ancestry did not report Sub-Saharan African origins and 5 patients self-reported such origins while they were not genetically clustered in the African cluster. Indeed, Cohen's kappa metric indicated a fair agreement (κ = 0.377). Considering low samples sizes, results may not be representative of the whole African / Sub-Saharan African MS population in France.

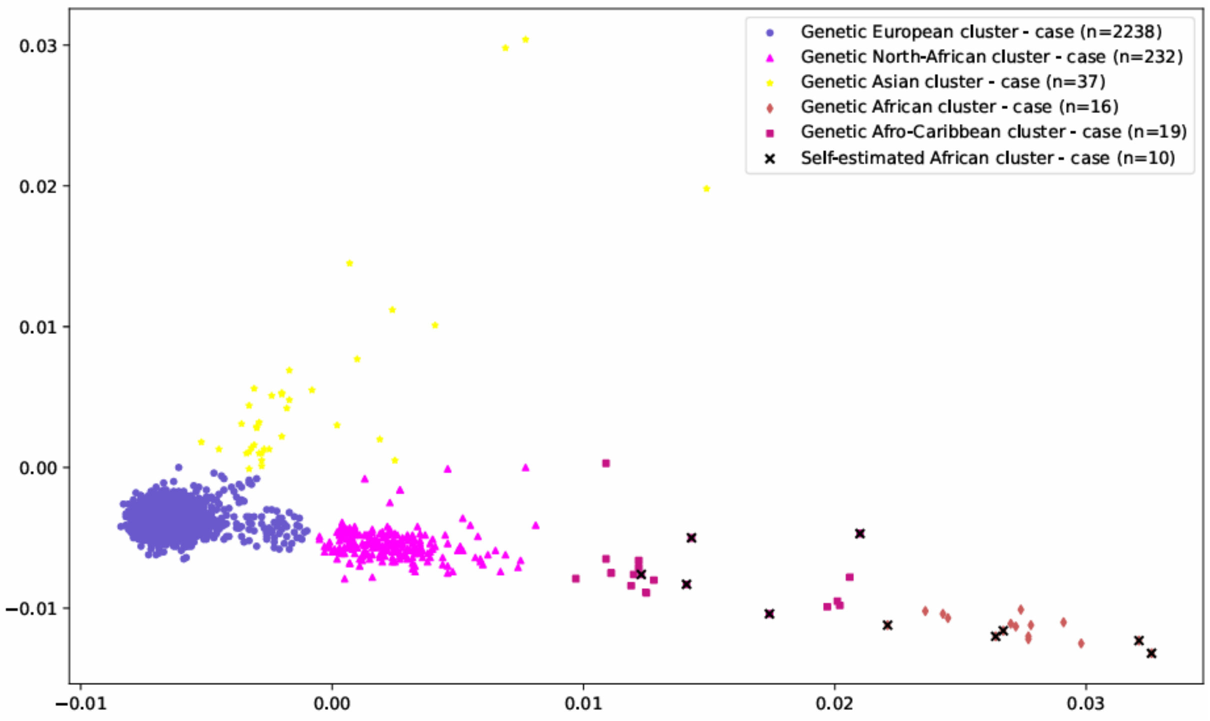


***Figure S6:*** *Self-reported Sub-Saharan African origins distribution displayed on the two first components of the PCA from ancestry analysis.*

Figure S6 shows the 5 self-reported Sub-Saharan origins patients genetically located in the Afro-Caribbean cluster, which may indicate a lower level of African ancestry. Note that on the other hand, not self-reporting as from Sub-Saharan African origins is in most cases an indicator of other genetic ancestral background.

*European ancestry*

European genetic clusters 1 and 7 were compared to European and French metropole self-reported origins. This cluster represent the group with the highest European ancestry.

| | **European or France metropole self-reported origins** | **Other self-reported origins** | **Total** |
|---|---|---|---|
| **European genetic ancestry** | 2,139 | 99 | 2,238 |
| **Other genetic ancestry** | 127 | 177 | 394 |
| **Total** | 2,266 | 276 | 2,542 |

***Table S13:*** *Contingency table of European (France metropole and European) self-reported individual origins' status compared to patients' European (cluster 1 and 2) genetic ancestry status. All expected frequencies are > 5 in all cells of the expected frequencies table (data not shown). We can use $X^2$ of independence test without Yate's correction.*

$X^2$ of independence test resulted in a significant association between European and Metropole French origins reporting and European genetic ancestry ($p < 0.001$). We found 127 patients from other genetic ancestry who self-reported European origins. Indeed, as demonstrated previously in North-African patients' analysis (Figure 3) and shown on Figure S5, an important proportion of patients from North-African genetic ancestry (127 patients) self-reported European origins. European self-reporting is associated with a moderate to substantial agreement with European genetic ancestry ($\kappa = 0.56$). However, data indicated a non-neglectable proportion of discordant pairs 8.89 %, mostly due to discrepancies with North-African patients (Figure S7).

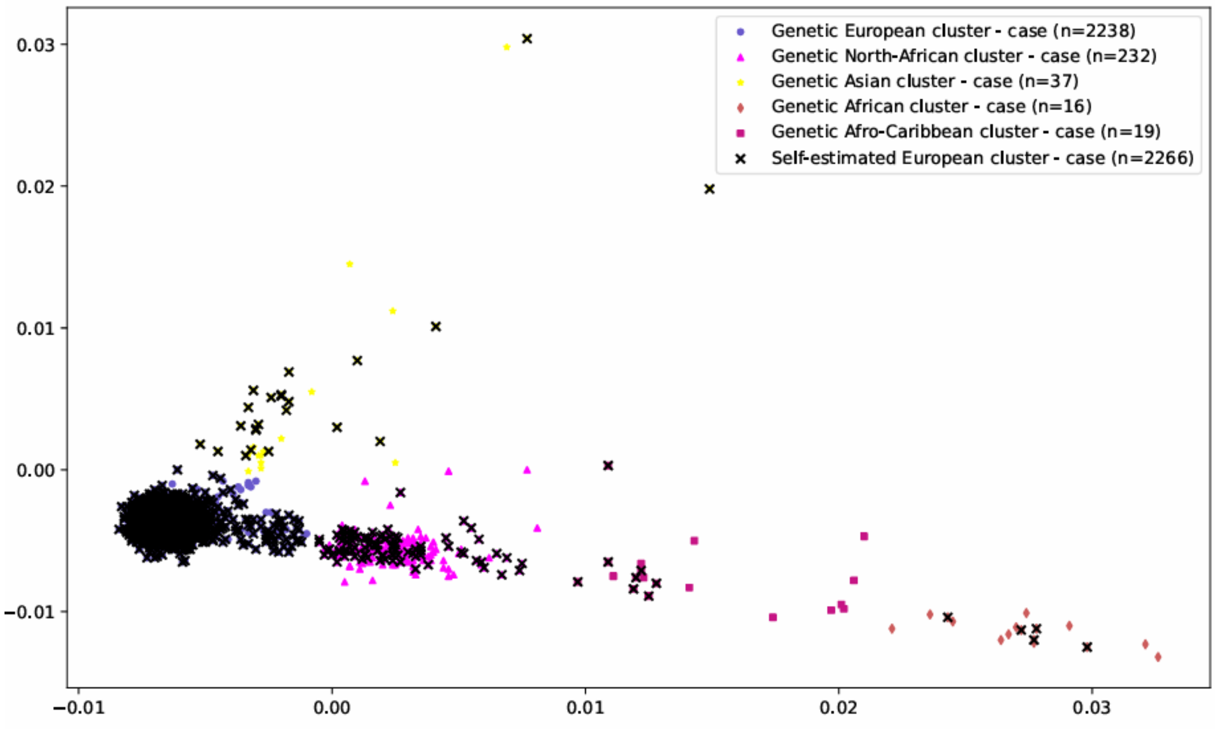


***Figure S7:*** *Self-reported France metropole and European origins distribution displayed on the two first components of the PCA from ancestry analysis.*

*Asian ancestry*

Cluster 4 and 6 from ancestry PCA were merged into one Asian genetic cluster, and compared to Asian self-reported origins (Table S14).

| | **South-Asian or East-Asian self-reported origins** | **Other self-reported origins** | **Total** |
|---|---|---|---|
| **Asian genetic ancestry** | 6 | 31 | 37 |
| **Other genetic ancestry** | 6 | 2,499 | 2,505 |
| **Total** | 12 | 2,530 | 2,542 |

***Table S14:*** *Contingency table of Asian self-reported individual origins' status compared to patients' Asian (cluster 4 and 6) genetic ancestry status. All expected frequencies are not > 5 in all cells of the expected frequencies table (data not shown). One cell (25%) shows very low sample size, indicating that we need to use an exact Fisher test.*

Fisher's exact test suggested a statistically significant association between Asian self-reported origins and genetic ancestry ($p < 0.01$). Out of a total of 37 patients from Asian ancestral cluster, only 6 (16.2 %) self-reported South or East-Asian origins. However, those extreme results may be due to the small sample size, yielding to low statistical power or biases. Another cause might be the overestimation of the Asian's genetic cluster size on the PCA. Moreover, biased may be induced by cultural sensitivities or even misunderstandings of questions in the case-report-form (CRF). Asian self-reported origins show fair agreement with Asian genetic ancestry (κ = 0.239). Figure S8 shows the six patients from other genetic ancestry who self-reported Asian origins. Those patients are located closer to the upper border of European clusters, of which some near the boundary with South-Asian cluster. This highlights the difficulties associated to genetic clustering methods in the context of admixed populations. Proportion of discordant pairs would not be meaningful in this configuration because of the limited sample size.

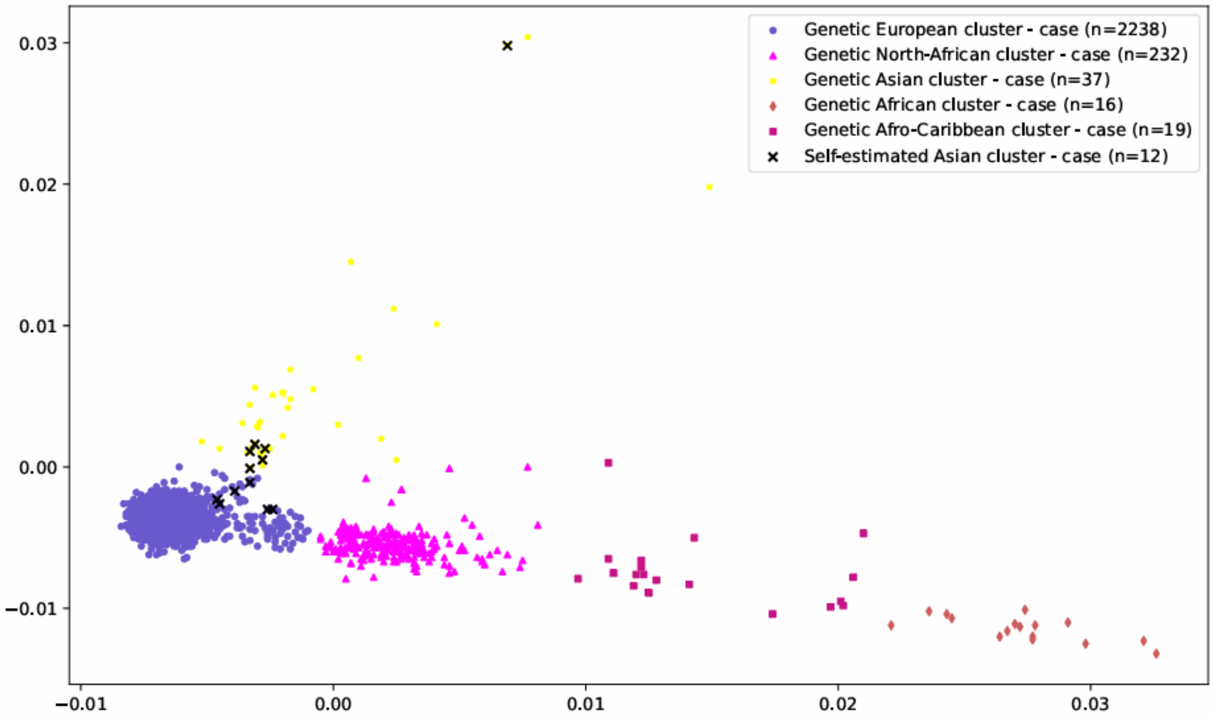

***Figure S8:*** *Self-reported Asian origins distribution displayed on the two first components of the PCA from ancestry analysis.*

*Afro-Caribbean ancestry*

Afro-Caribbean genetic ancestry (cluster 2) was compared to French overseas territories self-reported origins because of the large number of overseas territories located in regions close to central America and South-Africa (Table S15). This cluster represents an intermediate level of African ancestry, in between North-African and African clusters (respectively clusters 3 and 5).

| | Self-reported as Afro-Caribbean (French overseas territories) | Not self-reported as Afro-Caribbean (French overseas territories) | Total |
|---|---|---|---|
| **Afro-Caribbean genetic ancestry** | 2 | 17 | 19 |
| **Other genetic ancestry** | 12 | 2,511 | 2,523 |
| **Total** | 14 | 2,528 | 2,542 |

***Table S15:*** *Contingency table of Afro-Caribbean (French overseas territories) self-reported individual origins' status compared to patients' Afro-Caribbean (cluster 2) genetic ancestry status. All expected frequencies are not > 5 in all cells of the expected frequencies table (data not shown). One cell (25%) shows very low sample size, indicating that we need to use an exact Fisher test.*

Fisher's exact test resulted in a statistically significant association between Afro-Caribbean genetic ancestry and self-reported French overseas territories origins ($p < 0.05$). However, only two patients from Afro-Caribbean genetic ancestry self-reported originating from French overseas territories. Indeed, we found none to very slight agreement between self-report and genetic ancestry ($\kappa = 0.113$). Furthermore, patients not assigned to the Afro-Caribbean genetic cluster were six times more likely to self-report origins from French overseas territories ($n = 12$) than those in the Afro-Caribbean cluster ($n = 2$). This can be seen as the variety of French overseas territories gathering a wide range of ancestral backgrounds and cultures, yielding to potential biases. Additionally, considering the low sample size, it appears difficult to get trustful conclusions from those data. Moreover, not all patients from French overseas territories are from Afro-Caribbean ancestral background. This can be seen on Figure S9 with 7 patients genetically clustered between African and Asian clusters (South-Asian + East-Asian clusters) who self-reported French overseas territories origins. Four other patients self-reported those origins and were genetically associated to clusters with higher African ancestry.

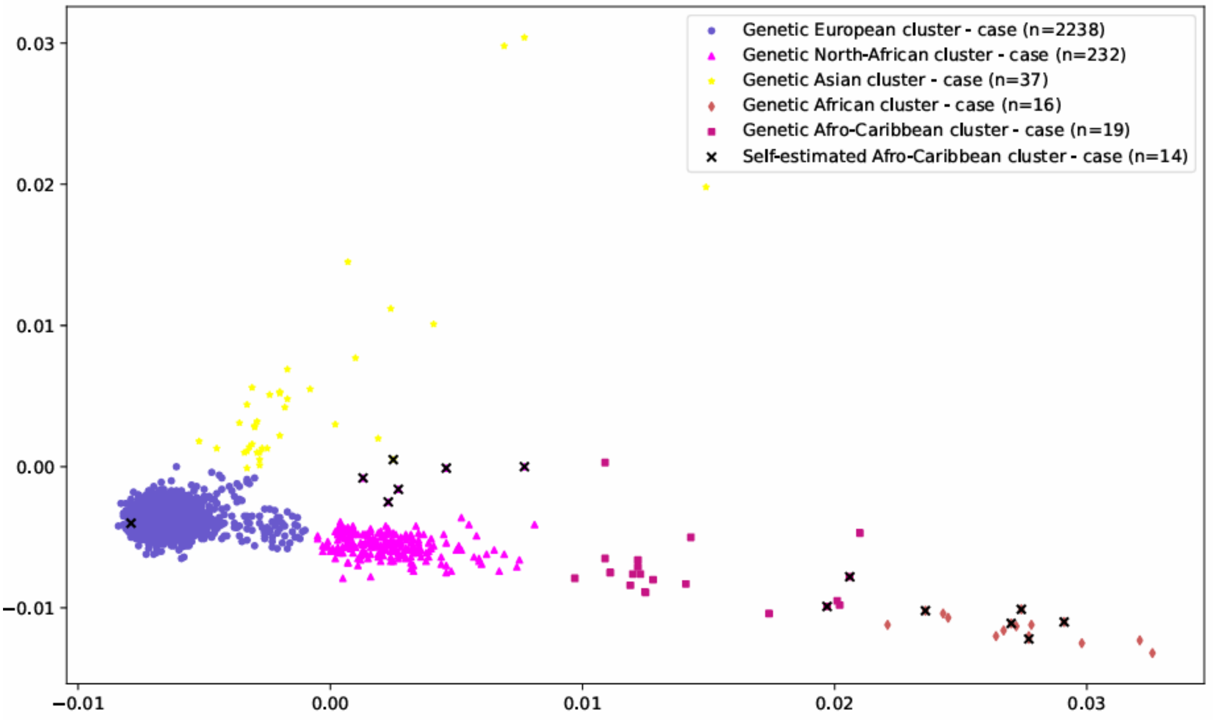


***Figure S9:*** *Self-reported French overseas territories origins distribution displayed on the two first components of the PCA from ancestry analysis.*

*Unknown and other origins*

Around 4.4% (113 / 2,542) of patients reported "unknown" or "others" origins making it impossible to compare with any defined genetic ancestral cluster. However, Figure S10 highlight this widely spread self-report status across all ancestral backgrounds, mostly in European and North-African clusters. Additionally, we detected a huge proportion of patients from Asian genetic ancestry self-reported "unknown" or "others' origins (relative to the number of patients genetically estimated as from Asian ancestry (n = 37) or self-reported from Asian origins (n = 12). Moreover, the important number of patients from North-African genetic cluster who self-reported "unknown" or "other" origins may reflect the social and cultural sensitivities associated to reporting North-African origins in France. Indeed, more than 20 patients from North-African genetic cluster self-reported “unknown” or “other” origins, representing more than 17 % of the 113 patients who self-reported “unknown” or “other” origins.

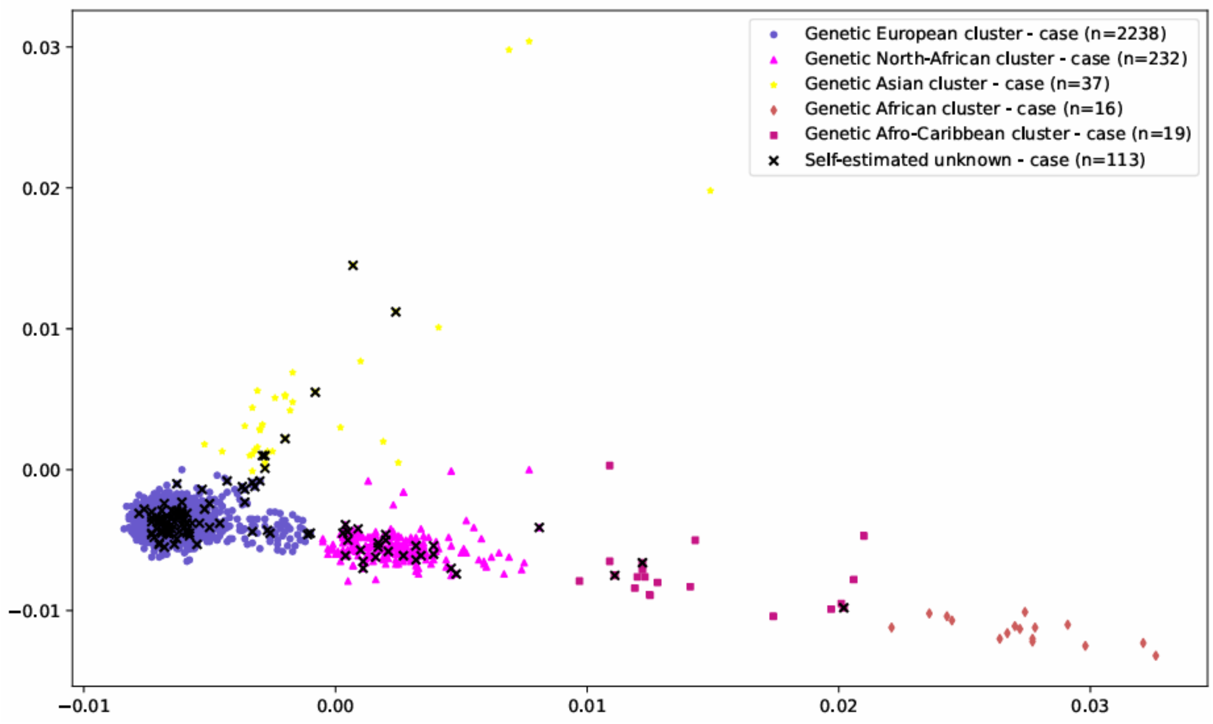


***Figure S10:*** *Self-reported unknown or “other” origins distribution displayed on the two first components of the PCA from ancestry analysis.*

### North-African patients and admixed genome proportions

We first focused on European genome proportions among North-African patients (Table S16).

| | North-African self-reported origins | Other self-reported origins |
|---|---|---|
| **North-African genetic ancestry** | A = 22.93 % ± 0.66 % | B = 23.86 % ± 0.91 % |
| **Other genetic ancestry** | C = 15.83 % ± 12.82 % | D = 2.54 % ± 0.37 % |

***Table S16****: Contingency table showing average African genome proportions regarding self-reported North-African origins status and genetic ancestry. Values are reported as mean ±* $MoE_{95\%}$ with $MoE95\% = 1.96 * \sigma/\sqrt{n}$ with $\sigma$ *the standard deviation and* $n$ *the total number of samples in the considered population.*

➔ *Question 1: Is African genome proportion significantly different between patients self-reported as North-African (A) and those who self-reported other origins (B) among patients from North-African genetic ancestry?*

Firsts, we needed to determine if we should perform a parametric student-t-test assuming normal distributions of samples or Mann-Whitney U test not assuming a priori normal data distribution. Thus, we performed statistical tests to assess for any significant deviation from a theoretical normal distribution. Visually, the distribution of African genome proportions of self-reported North-African origins patients among those from North-African genetic ancestry seems to follow a normal distribution (Figure S11).

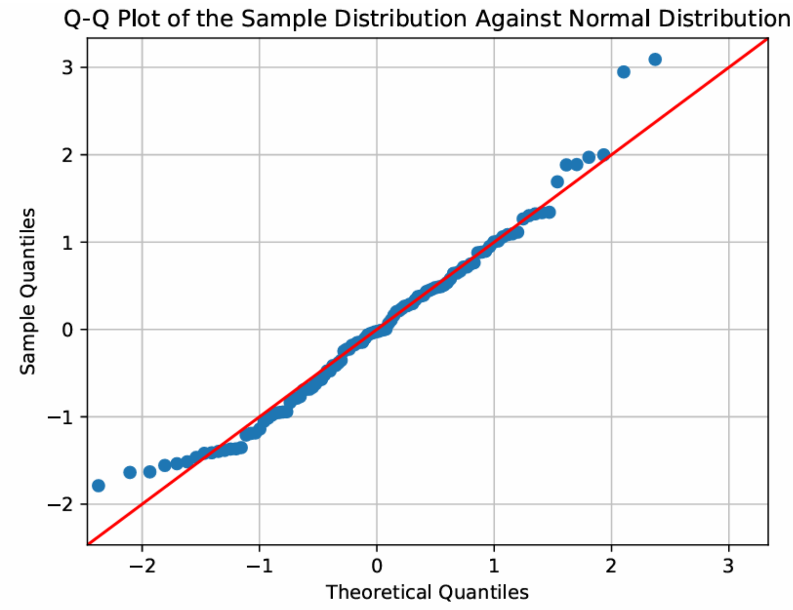


***Figure S11**: Quantile-quantile (QQ) plot showing African genome proportion distribution of patients' self-reported North-African origins (A) among patients from North-African genetic ancestry against theoretical quantiles of a normal distribution.*

We performed several normality tests for larger samples ($n_A > 50$): Kolmogorov-Smirnov one-sample test ($p = 0.92$), Anderson-Darling test ($p > 0.05$) and D'Agostino's K-squared test ($p = 0.129$). Those findings strongly indicate that distribution (A) does not significantly deviate from the normal theoretical distribution.

Likewise, we performed normality tests for distribution B corresponding to patients' self-reported other origins among patients from North-African genetic ancestry. Visually the distribution seems to deviate from theoretical normal distribution (Figure S12).

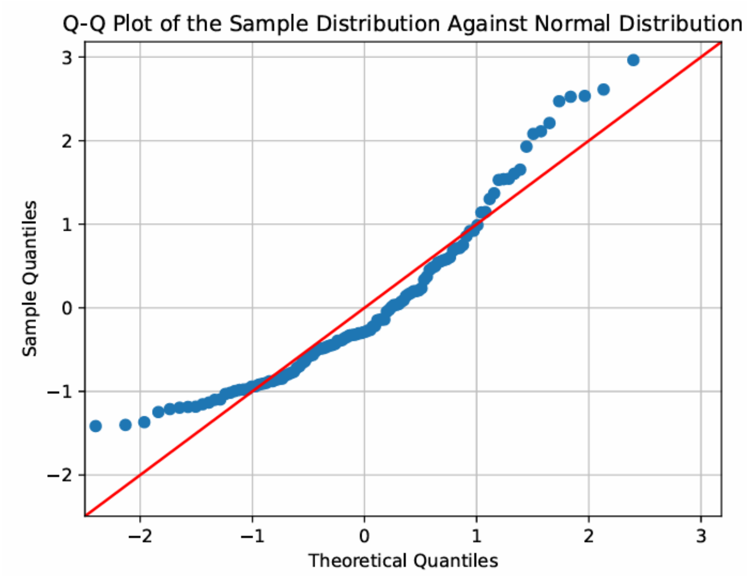


***Figure S12****: Quantile-quantile (QQ) plot showing African genome proportion distribution of patients' self-reported other origins (B) among patients from North-African genetic ancestry against theoretical quantiles of a normal distribution.*

We performed the same set of tests as for distribution A ($n_B > 50$): Kolmogorov-Smirnov one-sample test ($p < 0.05$), Anderson-Darling test ($p < 0.01$) and D'Agostino's K-squared test ($p < 0.01$). Those results suggest that distribution B significantly deviates from normal distribution. Thus, we chose to perform Mann-Whitney U test to compare distributions A and B.

Mann-Whitney U test resulted in no statistically significant African genome proportion differences in between patients self-reported as North-African and not self-reported from North-African origins among patients from North-African genetic ancestry ($p = 0.2106$). This suggests that self-report status regarding North-African origins is not associated with different African genome proportion when considering patients from North-African genetic ancestry.

➔ *Question 2: Is African genome proportion significantly different between patients self-reported as North-African (C) and those who self-reported other origins (D) among patients not from North-African genetic ancestry?*

Figure S13 shows substantial deviation of distribution C from normal theoretical distribution.

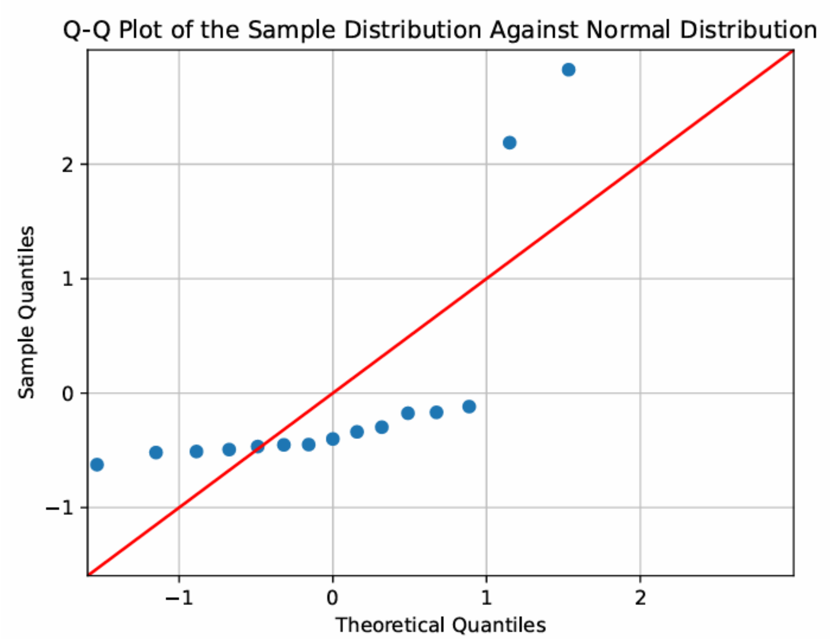


***Figure S13***: *Quantile-quantile (QQ) plot showing African genome proportion distribution of patients' self-reported North-African origins (C) among patients from other genetic ancestry against theoretical quantiles of a normal distribution.*

The sample size associated to distribution C is low ($n_C = 15 < 50$), making Shapiro-Wilk one-sample test more suitable to assess for normality. Results suggest that distribution C significantly deviates from normal distribution ($p < 1e-4$).

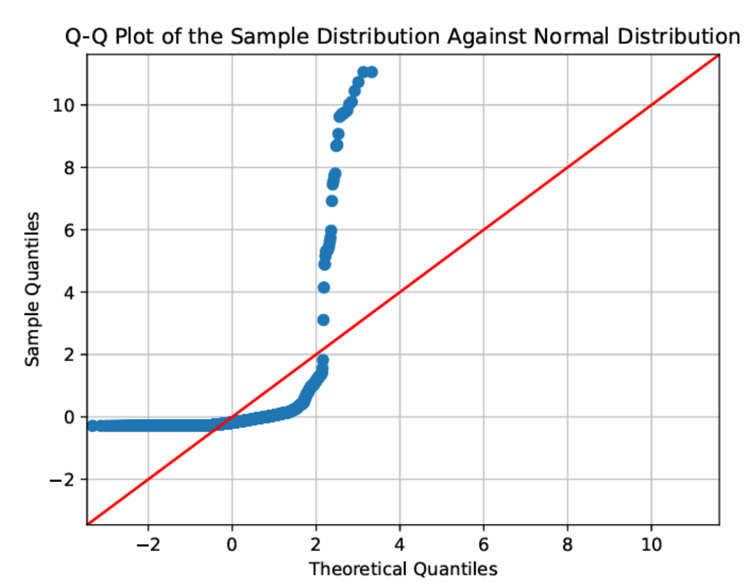


***Figure S14***: *Quantile-quantile (QQ) plot showing African genome proportion distribution of patients' self-reported other origins (D) among patients from other genetic ancestry against theoretical quantiles of a normal distribution.*

Figure S14 highlights the strong deviation of distribution D from normal theoretical distribution. We performed previously described normality tests for large sample sizes ($nD > 50$): Kolmogorov-Smirnov one-sample test ($p < 0.01$), Anderson-Darling test ($p < 0.01$) and D'Agostino's K-squared test ($p < 0.01$). Those findings indicate that distribution D does not follow a normal distribution and thus, we needed to use Mann-Whitney U test for non-parametric testing.

Mann-Whitney U test between distribution C and D resulted in a statistically significant difference ($p < 1e-4$) indicating that among patients not from North-African genetic ancestry, patients who self-reported North-African origins have a significantly higher proportion of African genome compared to patients who did not self-report such origins.

In addition, we implemented the same methodology for European genome proportions (Table S17).

| | **North-African self-reported origins** | **Other self-reported origins** |
|---|---|---|
| **North-African genetic ancestry** | A = 74.96% ± 0.69% | B = 74.20% ± 1.04% |
| **Other genetic ancestry** | C = 83.11% ± 12.67% | D = 95.73% ± 0.41% |

***Table S17:*** *Contingency table showing average European genome proportions regarding self-reported North-African origins and genetic ancestry status. Values are reported as mean* ± $MoE_{95\%}$ with $MoE95\% = 1.96 * \sigma/\sqrt{n}$ with $\sigma$ *the standard deviation and* $n$ *the total number of samples in the considered population.*

The following analysis of European genome proportions is included to enhance the reproducibility and confirm the robustness of findings from the African genome proportion analysis, given their largely complementary nature (neglectable East-Asian genome proportions). With African and European ancestries comprising the majority of the genome proportions in this cohort, the European analysis serves as a validation step to ensure consistency across ancestry-related findings (Table S17).

➔ *Question 3: Is European genome proportion significantly different between patients self-reported as North-African (A) and those who self-reported other origins (B) among patients from North-African genetic ancestry?*

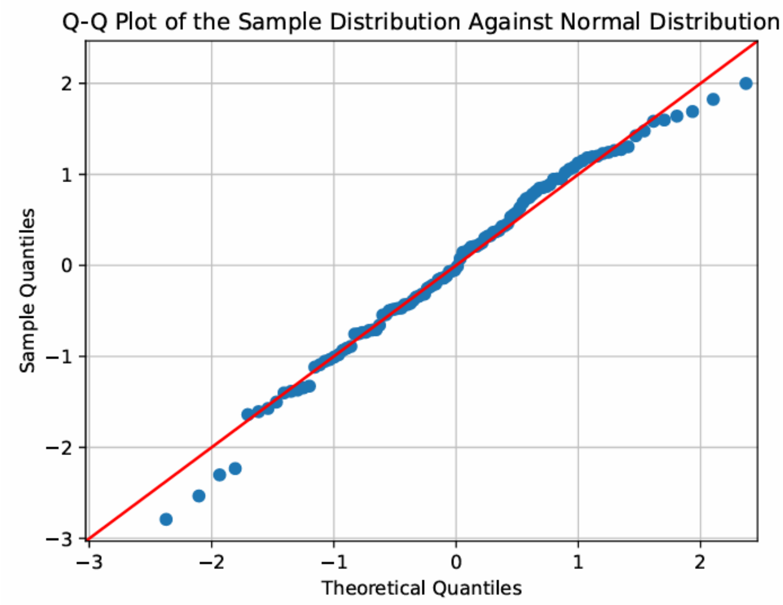


***Figure S15****: Quantile-quantile (QQ) plot showing European genome proportion distribution of patients' self-reported North-African origins (A) among patients from North-African genetic ancestry against theoretical quantiles of a normal distribution.*

Distribution A appears to not substantially deviate from normal distribution (Figure S 15). We performed normality testing for large samples like described previously ($n_A > 50$): Kolmogorov-Smirnov one-sample test ($p > 0.05$), Anderson-Darling test ($p > 0.15$) and D'Agostino's K-squared test ($p > 0.05$). Results indicate that distribution A does not significantly deviates from normal distribution.

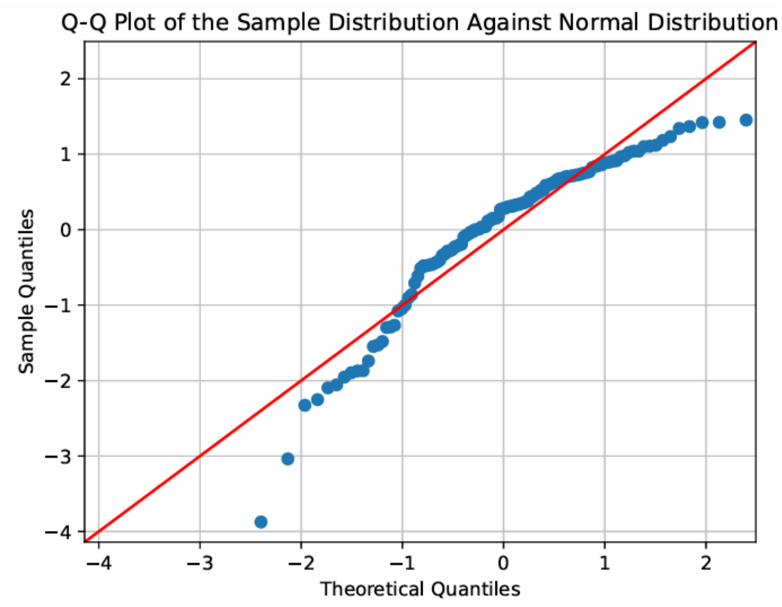


***Figure S16****: Quantile-quantile (QQ) plot showing European genome proportion distribution of patients' self-reported other origins (B) among patients from North-African genetic ancestry against theoretical quantiles of a normal distribution.*

On Figure S16, distribution B deviates from normal theoretical distribution. We performed normality testing for large samples like described previously ($n_B > 50$): Kolmogorov-Smirnov one-sample test ($p = 0.0505$; $p > 0.05$), Anderson-Darling test ($p < 0.01$) and D'Agostino's K-squared test ($p < 0.01$). Two normality tests out of three resulted in significant deviation of distribution B from normal distribution. Indeed, Kolmogorov-Smirnov test almost passed the 0.05 alpha threshold because of a lack of statistical power of the test compared to the two others[16].

Considering that both distributions (A and B) do not follow normal distribution, we performed Mann-Whitney U test between distributions A and B to test for European genome proportion differences. The test resulted in no significant differences regarding European genome proportions between both groups. This means that when an individual is genetically estimated as North-African, European genome proportions among North-African self-reported patients are not significantly different from the proportions among non-North-

African self-reported patients. This is consistent with the results obtained for African genome proportions.

➔ *Question 4: Is European genome proportion significantly different between patients self-reported as North-African (C) and patients who self-reported other origins (D) among patients not from North-African genetic ancestry?*

Figure S17 shows that distribution C of European genome proportions does not follow a normal theoretical distribution law. Considering the small sample size, we performed Shapiro-Wilk one-sample normality test ($n_C < 50$). The results indicate a significant deviation from normal theoretical distribution ($p < 0.01$).

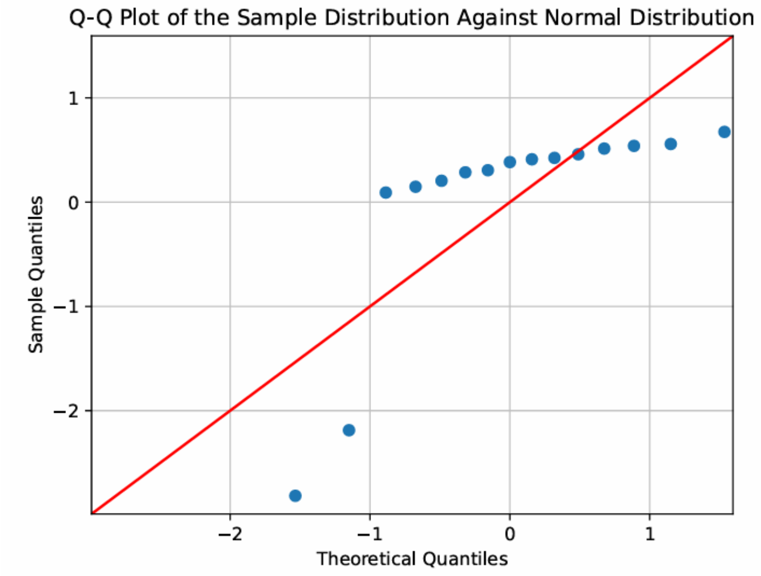


***Figure S17**: Quantile-quantile (QQ) plot showing European genome proportion distribution of patients' self-reported North-African origins (C) among patients from other genetic ancestry against theoretical quantiles of a normal distribution.*

Distribution D also seems to significantly deviate from normal distribution (Figure S18). We performed normality statistical testing for large samples following the same previously described process ($n_D > 50$): Kolmogorov-Smirnov one-sample test ($p < 0.01$), Anderson-Darling test ($p < 0.01$) and D'Agostino's K-squared test ($p < 0.01$). Findings indicate that distribution D is significantly deviating from normal distribution.

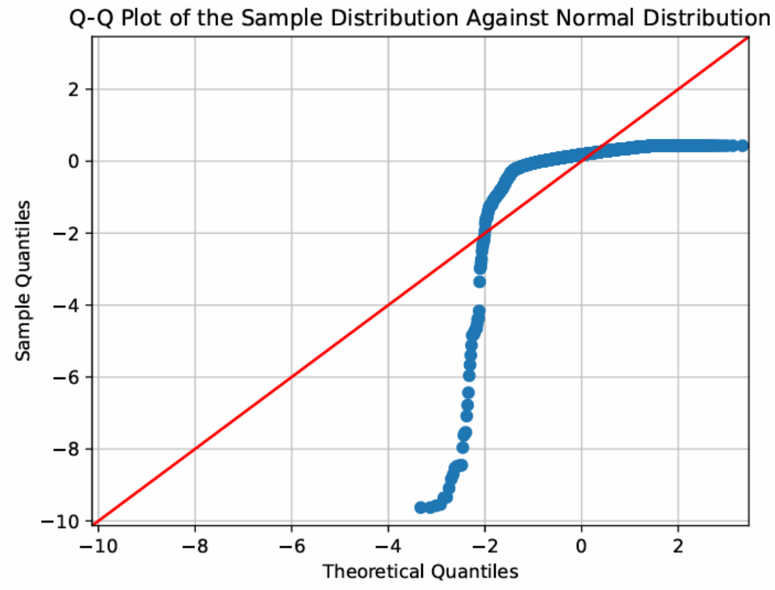


***Figure S18***: *Quantile-quantile (QQ) plot showing European genome proportion distribution of patients' self-reported other origins (D) among patients from other genetic ancestry against theoretical quantiles of a normal distribution.*

Thus, we used non-parametric Mann-Whitney U test to compare distributions C and D. We detected a significant difference regarding European genome proportions between patients' self-reported North-African origins and patients' self-reported other origins among patients from other genetic ancestry ($p < 1e-4$). Indeed, patients who self-reported other origins have a significantly higher proportion of European genome compared to patients who self-reported North-African origins, among patients from other genetic ancestry. This is consistent with previous results obtained on African genome proportions. We did not perform the analysis of Asian genome level since we considered the proportions as neglectable in the OFSEP-HD cohort.

### Analysis of admixed genome proportions of other genetic clusters

#### *African ancestry (cluster 5)*

On Table S18, self-reported Sub-Saharan-African origins appear to be associated with a higher African genome proportion (58.29 % ± 6.17 %) compared to patients who did not report such origins (3.92 % ± 0.31), among patients from other genetic ancestry. This Sub-Saharan-African self-identified origins of patients from other genetic ancestry might be explained by cultural affiliations, or because those patients did not show a sufficient proportion of African genome to be associated to the African genetic cluster (cluster 5). Thus, those patients might instead be genetically clustered in the Afro-Caribbean cluster (cluster 2).

| | **Self-reported Sub-Saharan-African origins** | **Other self-reported origins** |
|---|---|---|
| **African genetic ancestry** | 91.80 % ± 6.77 % | 88.47 % ± 2.8 % |
| **Other genetic ancestry** | 58.29 % ± 6.17 % | 3.92 % ± 0.31 % |

***Table S18:*** *Contingency table showing African genome proportions of OFSEP-HD patients with different status of African genetic ancestry and reported Sub-Saharan-African origins. Values are reported as mean* ± $MoE_{95\%}$ with $MoE95\% = 1.96 * \sigma/\sqrt{n}$ with $\sigma$ *the standard deviation and* $n$ *the total number of samples in the considered population.*

Table S19 illustrates the very low proportion of Asian genome among patients from African genetic ancestry. Because of the low sample size within the OFSEP-HD cohort, some results have larger $MoE_{95\%}$.

| | **Self-reported Sub-Saharan-African origins** | **Other self-reported origins** |
|---|---|---|
| **African genetic ancestry** | < 1e-5 % | 0.33 % ± 0.24 % |
| **Other genetic ancestry** | 3.85 % ± 3.90 % | 1.71 % ± 0.16 % |

***Table S19:*** *Contingency table showing Asian genome proportions of OFSEP-HD patients with different status of African genetic ancestry and reported Sub-Saharan-African origins. Values are reported as mean* ± $MoE_{95\%}$ with $MoE95\% = 1.96 * \sigma/\sqrt{n}$ with $\sigma$ *the standard deviation and* $n$ *the total number of samples in the considered population.*

Interestingly, patients from African ancestry show a non-neglectable proportion of European genome proportion in the range [1.43 %; 14.97 %] (Table S20). Consistent with previous results on African genome proportion showing higher values co-occurring with Sub-Saharan-African origins self-report, it appears that self-reporting Sub-Saharan-African origins is associated with lower proportions of European genome compared to patients who self-reported other origins.

| | **Self-reported Sub-Saharan-African origins** | **Other self-reported origins** |
|---|---|---|
| **African genetic ancestry** | 8.20 % ± 6.77 % | 11.20 % ± 2.75 % |
| **Other genetic ancestry** | 37.87 % ± 8.69 % | 94.36 % ± 0.35 % |

***Table S20:*** *Contingency table showing European genome proportions of OFSEP-HD patients with different status of African genetic ancestry and reported Sub-Saharan-African origins. Values are reported as mean* ± $MoE_{95\%}$ with $MoE95\% = 1.96 * \sigma/\sqrt{n}$ with $\sigma$ *the standard deviation and* $n$ *the total number of samples in the considered population.*

### *European ancestry (cluster 1 and 7)*

Table S21 clearly illustrate the high proportion of African genome among patients not from European genetic ancestry who self-reported French metropole / European origins (24.64 % ± 3.00 %). Those findings might reflect the French North-African population in France as described in the main paper.

| | **Self-reported European / French metropole origins** | **Other self-reported origins** |
|---|---|---|
| **European genetic ancestry** | 1.44 % ± 0.09 % | 3.03 % ± 0.69 % |
| **Other genetic ancestry** | 24.64 % ± 3.00 % | 28.85 % ± 2.85% |

***Table S21:*** *Contingency table showing African genome proportions of OFSEP-HD patients with different status of European genetic ancestry and reported European / French metropole origins. Values are reported as mean* ± $MoE_{95\%}$ with $MoE95\% = 1.96 * \sigma/\sqrt{n}$ with $\sigma$ *the standard deviation and* $n$ *the total number of samples in the considered population.*

European patients show neglectable proportions of Asian genome (Table S22). Nevertheless, those appear globally higher compared to African patients (c.f. above).

| | Self-reported European / French metropole origins | Other self-reported origins |
|---|---|---|
| **European genetic ancestry** | 1.31 % ± 0.06 % | 1.89 ± 0.45 % |
| **Other genetic ancestry** | 6.05 % ± 2.23 % | 3.32 % ± 1.36 % |

***Table S22:*** *Contingency table showing Asian genome proportions of OFSEP-HD patients with different status of European genetic ancestry and reported European / French metropole origins. Values are reported as mean* ± $MoE_{95\%}$ with $MoE95\% = 1.96 * \sigma/\sqrt{n}$ with $\sigma$ *the standard deviation and* $n$ *the total number of samples in the considered population.*

The large majority of patients in the OFSEP-HD cohort are from European genetic ancestral background, explaining high proportions of European genome among all self-report sub-categories of European patients (Table S23).

| | Self-reported European / French metropole origins | Other self-reported origins |
|---|---|---|
| **European genetic ancestry** | 97.25 % ± 0.10 % | 95.07 ± 0.90 % |
| **Other genetic ancestry** | 69.31 % ± 3.01 % | 67.83 % ± 2.81 % |

***Table S23:*** *Contingency table showing European genome proportions of OFSEP-HD patients with different status of European genetic ancestry and reported European / French metropole origins. Values are reported as mean* ± $MoE_{95\%}$ with $MoE95\% = 1.96 * \sigma/\sqrt{n}$ with $\sigma$ *the standard deviation and* $n$ *the total number of samples in the considered population.*

*Afro-Caribbean ancestry (cluster 2)*

Compared to African ancestry (cluster 5), Afro-Caribbean ancestry is clearly an admixed population with intermediate levels of African genome proportions, presenting a higher proportion of African ancestral background compared to North-African patients (cluster 3) and a lower one than African patients (Table S24). This aligns with our expectations and strengthen the methodology consisting of mapping French overseas territories origins with Afro-Caribbean ancestry (cluster 2).

| | Self-reported French overseas territories origins | Other self-reported origins |
|---|---|---|
| **Afro-Caribbean genetic ancestry** | 69.48 % ± 1.69 % | 53.72 % ± 4.36 % |
| **Other genetic ancestry** | 47.48 % ± 19.39 % | 3.98 % ± 0.35 % |

***Table S24:*** *Contingency table showing African genome proportions of OFSEP-HD patients with different status of Afro-Caribbean genetic ancestry and reported French overseas territories origins. Values are reported as mean* ± $MoE_{95\%}$ with $MoE95\% = 1.96 * \sigma/\sqrt{n}$ with $\sigma$ *the standard deviation and* $n$ *the total number of samples in the considered population.*

Afro-Caribbean patients show residual proportions of Asian genome (Table S25) comparable to Asian genome proportion in patients from African ancestry (cluster 5).

| | **Self-reported French overseas territories origins** | **Other self-reported origins** |
|---|---|---|
| **Afro-Caribbean genetic ancestry** | 2.01 % ± 2.78 % | 2.79 % ± 2.35 % |
| **Other genetic ancestry** | 6.54 % ± 3.72 % | 1.68 % ± 0.16 % |

***Table S25:*** *Contingency table showing East-Asian genome proportions OFSEP-HD patients with different status of Afro-Caribbean genetic ancestry and reported French overseas territories origins. Values are reported as mean* ± $MoE_{95\%}$ with $MoE95\% = 1.96 * \sigma/\sqrt{n}$ with $\sigma$ *the standard deviation and* $n$ *the total number of samples in the considered population.*

Table S26 illustrates the admixture nature of Afro-Caribbean ancestry, notably with French European ancestry, highlighted by significant proportion of European genome among all sub categories of Afro-Caribbean patients.

| | **Self-reported French overseas territories origins** | **Other self-reported origins** |
|---|---|---|
| **Afro-Caribbean genetic ancestry** | 28.51 % ± 4.47 % | 43.49 % ± 4.54 % |
| **Other genetic ancestry** | 45.98 % ± 17.16 % | 94.34 % ± 0.39 % |

***Table S26:*** *Contingency table showing European genome proportions OFSEP-HD patients with different status of Afro-Caribbean genetic ancestry and reported French overseas territories origins. Values are reported as mean* ± $MoE_{95\%}$ with $MoE95\% = 1.96 * \sigma/\sqrt{n}$ with $\sigma$ *the standard deviation and* $n$ *the total number of samples in the considered population.*

*Asian ancestry (cluster 4 and 6)*

Self-reported and genetically estimated Asian patients show residual proportions of African genome proportions (Table S27). We noted the high variability in genome proportions due to insufficient sample size associated to Asian ancestry (Table S28). European genome proportion is substantially high among reported or genetically estimated Asian patients (Table

S29). Indeed, this subpopulation surprisingly exhibits more than two times higher European genome proportions compared to proportions of Asian genome, with estimated European proportion in the range [45.56 %; 93.40 %]. These findings might be explained by the strong European-East-Asian admixed nature of South-Asian populations (included in Asian genetic cluster), *further* increased by the overrepresentation of South-Asian patients in the Asian genetic cluster compared to the number of patients from East-Asian genetic ancestry.

| | Self-reported Asian origins | Other self-reported origins |
|---|---|---|
| **Asian genetic ancestry** | 3.73 % ± 1.45 % | 5.98 % ± 2.23 % |
| **Other genetic ancestry** | 6.37 % ± 2.06 % | 4.55 % ± 0.42 % |

***Table S27:*** *Contingency table showing African genome proportions OFSEP-HD patients with different status of Asian genetic ancestry and reported Asian origins. Values are reported as mean* ± $MoE_{95\%}$ with $MoE95\% = 1.96 * \sigma/\sqrt{n}$ with $\sigma$ *the standard deviation and* $n$ *the total number of samples in the considered population.*

| | Self-reported Asian origins | Other self-reported origins |
|---|---|---|
| **Asian genetic ancestry** | 26.79 % ± 25.27 % | 25.23 % ± 6.42 % |
| **Other genetic ancestry** | 4.57 % ± 1.39 % | 1.35 % ± 0.06 % |

***Table S28:*** *Contingency table showing East-Asian genome proportions OFSEP-HD patients with different status of Asian genetic ancestry and reported Asian origins. Values are reported as mean* ± $MoE_{95\%}$ with $MoE95\% = 1.96 * \sigma/\sqrt{n}$ with $\sigma$ *the standard deviation and* $n$ *the total number of samples in the considered population.*

| | Self-reported Asian origins | Other self-reported origins |
|---|---|---|
| **Asian genetic ancestry** | 69.48 % ± 23.92 % | 68.79 % ± 7.18 % |
| **Other genetic ancestry** | 89.05 % ± 2.24 % | 94.09 % ± 0.43 % |

***Table S29:*** *Contingency table showing European genome proportions OFSEP-HD patients with different status of Asian genetic ancestry and reported Asian origins. Values are reported as mean* ± $MoE_{95\%}$ with $MoE95\% = 1.96 * \sigma/\sqrt{n}$ with $\sigma$ *the standard deviation and* $n$ *the total number of samples in the considered population.*

**Parents' North-African origins: a better proxy of patients' genetic ancestry?**

- Details on independent data configuration analysis

We found no statistically significant association between parents' North-African origins and patients' genetic ancestry in this configuration (p = 1). This indicates that neither fathers nor mothers show a better agreement with patients' genetic ancestry (Table S30). This highlights that relying solely on fathers' or mothers' self-reported North-African origins should not be used as a proxy to assess for the genetic ancestry patients, especially in the context of the French MS population.

| | Father self-reported North-African origins | Mother self-reported North-African origins | Total |
|---|---|---|---|
| **North-African genetic ancestry** | 11 | 4 | 15 |
| **Other genetic ancestry** | 58 | 26 | 84 |
| **Total** | 69 | 30 | 99 |

***Table S30:*** *Contingency table of parents' North-African origins compared to patients' North-African genetic ancestry in cases of discrepancy between parents' report regarding patient's North-African genetic ancestry. Father self-reported North-African origins: only the father self-reported as North-African. Mother self-reported North-African origins: only the mother self-reported as North-African. All expected frequencies are not > 5 in all cells of the expected frequencies table (data not shown). One cell (25%) shows very low sample size, indicating that we need to use an exact Fisher test to compare parents' North-African origins and patients' genetic ancestry.*

- Details on paired data configuration analysis

| | Father self-reported North-African origins | Father self-reported other origins | Mother self-reported North-African origins | Mother self-reported other origins | Both self-reported North-African origins | Both self-reported other origins |
|---|---|---|---|---|---|---|
| **North-African genetic ancestry** | 183 | 49 | 176 | 56 | 172 | 45 |
| **Other genetic ancestry** | 89 | 2,221 | 57 | 2,253 | 31 | 2,195 |

***Table S31****: Paired contingency tables showing parents' North-African origins status compared to patients' genetic ancestry.*

| | **Father self-reported North-African origins** | **Father self-reported other origins** |
|---|---|---|
| **North-African genetic ancestry** | 183 | 49 |
| **Other genetic ancestry** | 89 | 2,221 |

***Table S32**: Paired contingency tables showing fathers' self-reported North-African origins status compared to patients' genetic ancestry.*

| | **Mother self-reported North-African origins** | **Mother self-reported other origins** |
|---|---|---|
| **North-African genetic ancestry** | 176 | 56 |
| **Other genetic ancestry** | 57 | 2,253 |

***Table S33**: Paired contingency tables showing mothers' self-reported North-African origins status compared to patients' genetic ancestry.*

| | **Both self-reported North-African origins** | **Both self-reported other origins** |
|---|---|---|
| **North-African genetic ancestry** | 172 | 45 |
| **Not North-African genetic ancestry** | 31 | 2,195 |

***Table S34:** Paired contingency tables showing parents' North-African origins status compared to patients' genetic ancestry in cases where both parents have the same origins.*

Fathers' only % discordant pairs: (4 + 58) / (11 + 4 + 58 + 26) * 100 = 62.62 %

Mothers' only % discordant pairs: (11 + 26) / (11 + 4 + 58 + 26) * 100 = 37.37 %

Common parents' origins % discordant pairs: (45 + 31) / (172 + 45 + 31 + 2,195) * 100 = 3.11 %

### Other parents' origins and genetic ancestries

#### *African ancestry (cluster 5)*

One of the expected frequencies is largely below 5, representing 25% of the cells, meaning that we needed to use Fisher's exact test to perform the analysis (data not shown). We found a statistically significant association between parents' Sub-Saharan-African origins status and African ancestry of the child ($p < 0.001$) but only when both parents have the same origins, especially driven by cases where both parents did not report Sub-Saharan-African origins and patients were not from African ancestry (Table S35). We evaluated a fair agreement with Cohen's Kappa value $\kappa = 0.30 \pm 0.003$ when both parents exhibited common self-report of Sub-Saharan-African origins and an overall low proportion of discordant pairs (1.18 %), mainly due to small sample size associated to Sub-Saharan-African patients in the cohort. However, it seems important to note that approximately one half of cases where both parents' Sub-Saharan-African origins are false positive in regards to patients' African genetic ancestry. In addition, the low sample size may have led to potential biases and weak statistical power during this analysis.

| | Father self-reported as Sub-Saharan-African | Mother self-reported as Sub-Saharan-African | Both self-reported as Sub-Saharan-African | None self-reported as Sub-Saharan-African | Total |
|---|---|---|---|---|---|
| **African genetic ancestry** | 1 | 1 | 6 | 8 | 16 |
| **Not African genetic ancestry** | 9 | 6 | 7 | 2,504 | 2,526 |
| **Total** | 10 | 7 | 13 | 2,512 | 2,542 |

***Table S35:*** *Contingency table presenting parents' Sub-Saharan-African origins in relation to patients' African genetic ancestry. Data are shown in independent manner. Father self-reported as Sub-Saharan-African: only the father self-reported as Sub-Saharan-African. Mother self-reported as Sub-Saharan-African: only the mother self-reported as Sub-Saharan-African.*

*European ancestry (clusters 1 and 7)*

European clusters totalize 2,238 patients representing more than 88 % of the whole OFSEP-HD cohort, which is consistent with the diversity within French MS population. Nonetheless, results from $X^2$ test without need of Yate's correction (no expected frequencies is $\leq 5$; data not shown) indicated a statistically significant association between parents' European / French metropole origins and patients' European genetic ancestry ($p < 0.001$). This is further highlighted by the measured substantial agreement for fathers ($\kappa$ = 0.636 ± 0.003) and mothers ($\kappa$ = 0.669 ± 0.003) self-reported origins with patients' European genetic ancestry. Additionally, we found a lower but still moderate agreement between common parents' origins and patients' European genetic ancestry ($\kappa$ = 0.576 ± 0.001).

| | **Father self-reported as French metropole / European** | **Mother self-reported as French metropole / European** | **Both self-reported as French metropole / European** | **None self-reported as French metropole / European** | **Total** |
|---|---|---|---|---|---|
| **European genetic ancestry** | 39 | 94 | 1,982 | 123 | 2,238 |
| **Other genetic ancestry** | 13 | 28 | 16 | 247 | 304 |
| **Total** | 52 | 122 | 1,998 | 370 | 2,542 |

***Table S36:*** *Contingency table presenting parents' French metropolitan/European origins in relation to patients' European genetic ancestry. Data are shown in independent manner. Father self-reported as French metropole / European: only the father self-reported as French metropole / European. Mother self-reported as French metropole / European: only the mother self-reported as French metropole / European.*

*Afro-Caribbean ancestry (cluster 2)*

Afro-Caribbean ancestry and French overseas territories origins show very low sample size, with only 19 patients from Afro-Caribbean genetic ancestry (Table S37). Nevertheless, more than 89 % (17/19) of parents or patients from Afro-Caribbean genetic ancestry did not report French overseas territories origins. This particularly weak agreement even in common parents' origins configuration ($\kappa$ = 0.085 ± 0.0026) may either indicate that sample size is too low to get any insights, or that mapping Afro-Caribbean ancestry (cluster 2) with French overseas territories origins was not the best fit. Indeed, distinct classification of French overseas territories origins regarding ancestral background is challenging, given the diverse and geographically dispersed locations of these territories across the globe.

| | **Father self-reported as French overseas territories** | **Mother self-reported as French overseas territories** | **Both self-reported as French overseas territories** | **None self-reported as French overseas territories** | **Total** |
|---|---|---|---|---|---|
| **Afro-Caribbean genetic ancestry** | 0 | 0 | 2 | 17 | 19 |
| **Other genetic ancestry** | 7 | 4 | 10 | 2,502 | 2,523 |
| **Total** | 7 | 4 | 12 | 2,519 | 2,542 |

***Table S37:*** *Contingency table presenting parents' French overseas territories origins in relation to patients' Afro-Caribbean genetic ancestry. Data are shown in independent manner. Father self-reported as with French overseas territories origins: only the father self-reported as with French overseas territories origins. Mother self-reported as with French overseas territories origins: only the mother self-reported as with French overseas territories origins.*

### *Asian ancestry (clusters 4 and 6)*

Parents' Asian origins demonstrated a significant association with Asian genetic ancestry of their child ($p < 0.001$) (Table S38). We chose to perform Fisher's exact test because 50 % of the cells (4 / 8) values below 5 (data not shown). However, agreement appears similarly from fair to moderate for all parents' categories, when taken individually ($\kappa_{Father}$= 0.415 ± 0.045; $\kappa_{Mother}$= 0.427 ± 0.046) or for common self-reported origins ($\kappa_{Common}$= 0.387 ± 0.0026). Indeed, out of the 37 patients from Asian genetic ancestry, only 16 have at least one parent self-reporting Asian origins, representing less than one half of patients from Asian genetic ancestral background. We believe that sample size is too small in this case to get any meaningful insight comparing Asian ancestry and reported origins.

| | Father self-reported as Asian | Mother self-reported as Asian | Both self-reported as Asian | None self-reported as Asian | Total |
|---|---|---|---|---|---|
| **Asian genetic ancestry** | 3 | 3 | 10 | 21 | 37 |
| **Not Asian genetic ancestry** | 4 | 2 | 7 | 2,492 | 2,505 |
| **Total** | 7 | 5 | 17 | 2,513 | 2,542 |

***Table S38:*** *Contingency table presenting parents' Asian origins in relation to patients' Asian genetic ancestry. Data are shown in independent manner. Father self-reported as Asian: only the father self-reported as Asian. Mother self-reported as Asian: only the mother self-reported as Asian.*

### Details on HLA alleles imputation

The metric returned by HIBAG imputation software is a post probability representing how much we can trust results. In practice, a post probability above 0.5 can be trusted, and represents a prediction accuracy of 80%.

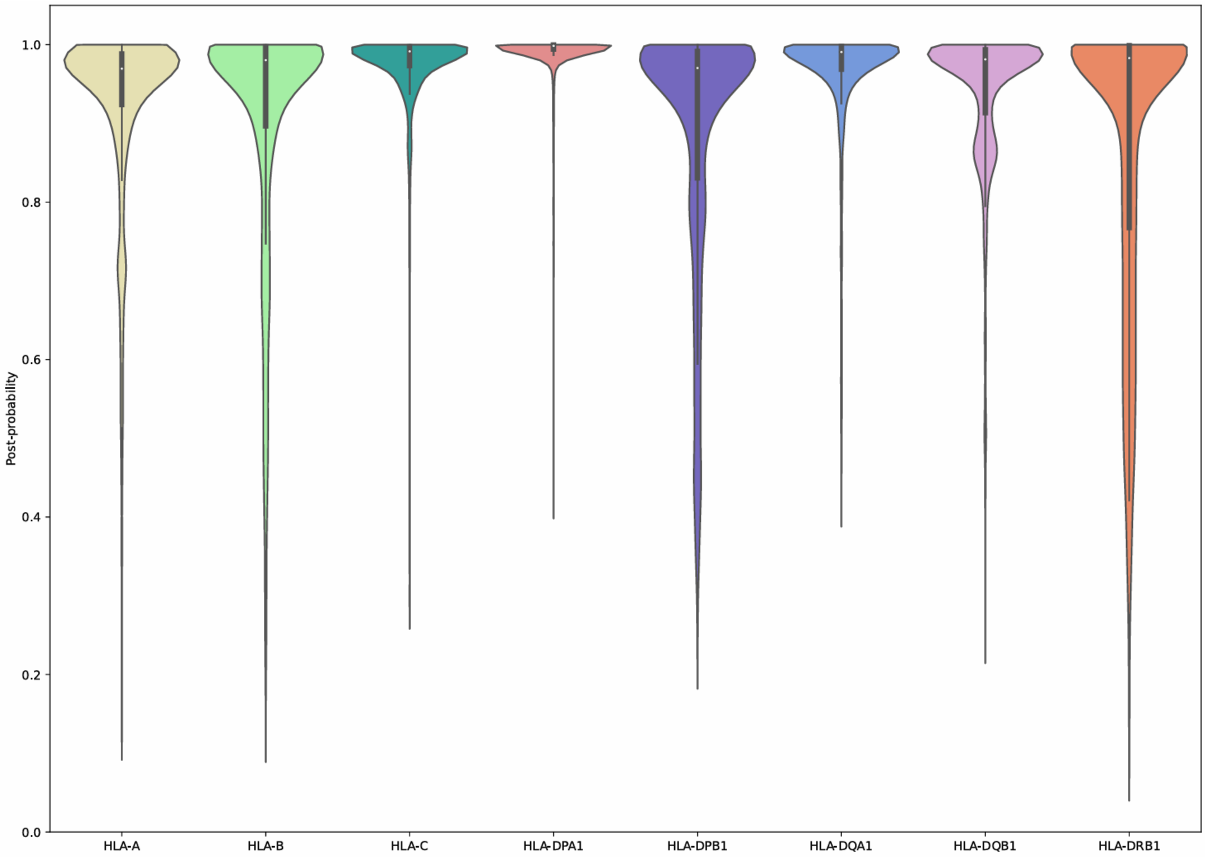


***Figure S19:*** *Distribution of imputation post-probabilities for each of the considered HLA class I (HLA-A, HLA-B, HLA-C) and class II (HLA-DPA1, HLA-DPB1, HLA-DQA1, HLA-DQB1, HLA-DRB1) genes.*

| HLA gene | Mean | SD | Min | Max | Q1 | Q3 |
|---|---|---|---|---|---|---|
| **A** | 92.5 % | **19.4 %** | 14.0 % | 99.9 % | 92.4 % | 98.8 % |
| **B** | 90.4 % | 11.5 % | 15.5 % | **100 %** | 89.7 % | 99.7 % |
| **C** | 97.1 % | 15.9 % | 28.6 % | 99.9 % | **97.3 %** | 99.7 % |
| **DQA1** | 96.7 % | 6.6 % | **41.7 %** | 99.9 % | 96.9 % | 99.8 % |
| **DPB1** | 88.5 % | 4.2 % | 25.2 % | 99.9 % | 83.1 % | 99.1 % |
| **DPA1** | **98.9 %** | 16.7 % | 41.6 % | 99.9 % | 99.4 % | **99.9 %** |

| | | | | | | |
|---|---|---|---|---|---|---|
| **DQB1** | 94.1 % | 7.0 % | 25.2 % | 99.9 % | 91.4 % | 99.4 % |
| **DRB1** | **86.6 %** | 9.0 % | 12.1 % | **100 %** | 76.8 % | **99.9 %** |

***Table S39:*** *Descriptive statistics of alleles imputation quality for each of considered HLA class I and II genes.*

### Details on HLA haplotypes imputation

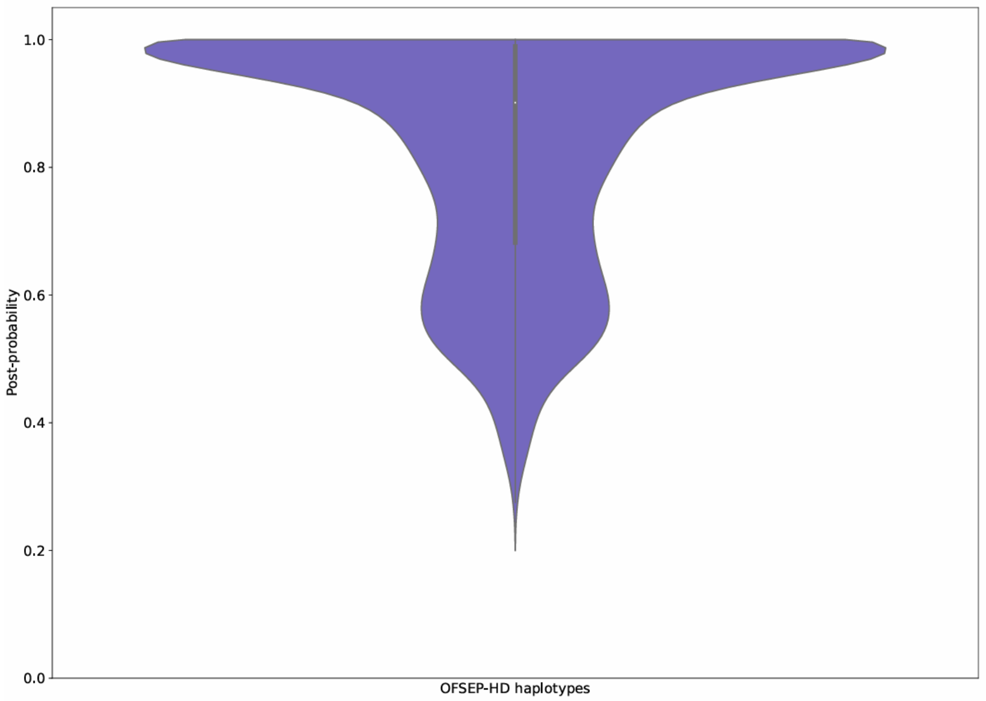


***Figure S20:*** *Distribution of imputation post-probabilities for all imputed haplotypes including loci HLA-A, HLA -B, HLA-C, HLA-DR and HLA-DQ.*

| | Mean | SD | Min | Max | Q1 | Q3 |
|---|---|---|---|---|---|---|
| **Haplotype imputation post-probabilities** | 82.9 % | 18.1% | 27.5% | 100% | 68.2% | 98.9% |

***Table S40:*** *Descriptive statistics of haplotypes imputation quality.*

### Computing polygenic risk scores

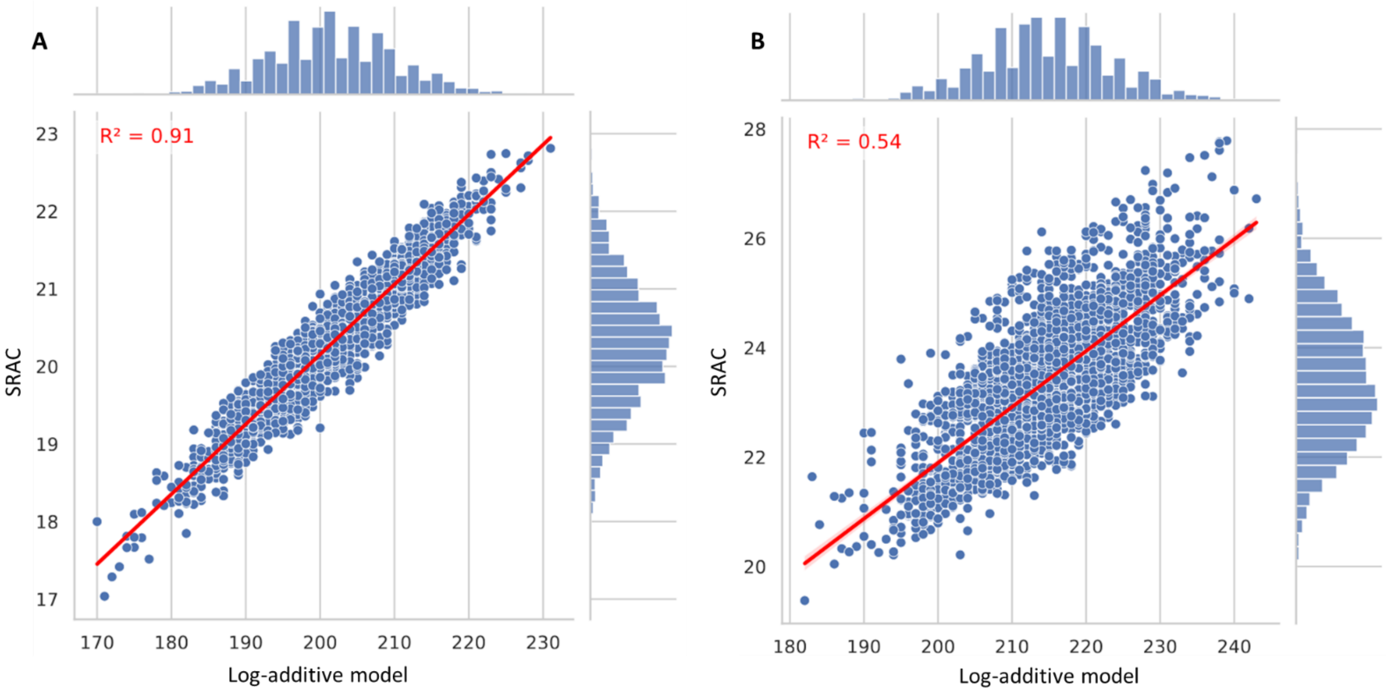


***Figure S21:*** *(A) Correlation plot between SRAC (Sum of Risk Alleles Count) and log-additive model using 190 non-MHC MS susceptibility SNPs. (B) Correlation plot between SRAC and log-additive model using 210 MS susceptibility associated variants, including the available 13 MHC region SNPs and 7 HLA alleles.*

| | | Mean ± MoE | SD | Min | Max | Q1 | Q3 |
|---|---|---|---|---|---|---|---|
| OFSEP-HD (n = 2542) | Log-additive | 20.29 ± 0.03 | 0.82 | 17.04 | 22.81 | 19.77 | 20.82 |
| | SRAC | 201.45 ± 0.34 | 8.66 | 170 | 231 | 196 | 207 |
| | Log-additive (+ MHC-variants) | 23.38 ± 0.05 | 1.23 | 19.38 | 27.78 | 22.50 | 24.21 |
| | SRAC (+ MHC variants) | 214.52 ± 0.34 | 8.85 | 182 | 243 | 209 | 220 |
| North-African genetic ancestry cluster (n = 232) | Log-additive | 19.99 ± 0.10 | 0.80 | 17.79 | 22.74 | 17.97 | 18.19 |
| | SRAC | 198.54 ± 1.09 | 8.44 | 176 | 225 | 178.31 | 180.73 |
| | Log-additive (+ MHC-variants) | 22.98 ± 0.14 | 1.06 | 20.53 | 25.78 | 20.57 | 20.64 |
| | SRAC (+ MHC variants) | 211.91 ± 1.08 | 8.40 | 191 | 238 | 192.73 | 194.73 |
| European genetic ancestry cluster (n = 2177) | Log-additive | 20.32 ± 0.03 | 0.81 | 17.04 | 22.81 | 17.80 | 18.32 |
| | SRAC | 201.79 ± 0.36 | 8.65 | 170 | 231 | 175 | 181 |
| | Log-additive (+ MHC-variants) | 23.45 ± 0.05 | 1.23 | 20.04 | 27.78 | 20.36 | 20.78 |
| | SRAC (+ MHC variants) | 214.85 ± 0.37 | 8.85 | 183 | 243 | 187.44 | 193 |

***Table S41:*** *Polygenic Risk Scores distribution characteristics computed on the whole population of 2542 considered OFSEP-HD patients as well as on North-African and European genetic ancestral clusters. SD: Standard Deviation. Min: Minimum. Max: Maximum. Q1: 25th percentile. Q3: 75th percentile. Log-additive model is the model weighted by variants effect size.* $MoE_{95\%} = 1.96 * \frac{\sigma}{\sqrt{n}}$, *$\sigma$ the standard deviation (SD) and $n$ the total number of samples in the considered population.*

To compare North-African and European ancestry score distributions, we used Non-parametric Mann-Whitney U tests since this is the standard test when comparing PRS distributions in the literature (Figure S22)[17,18].

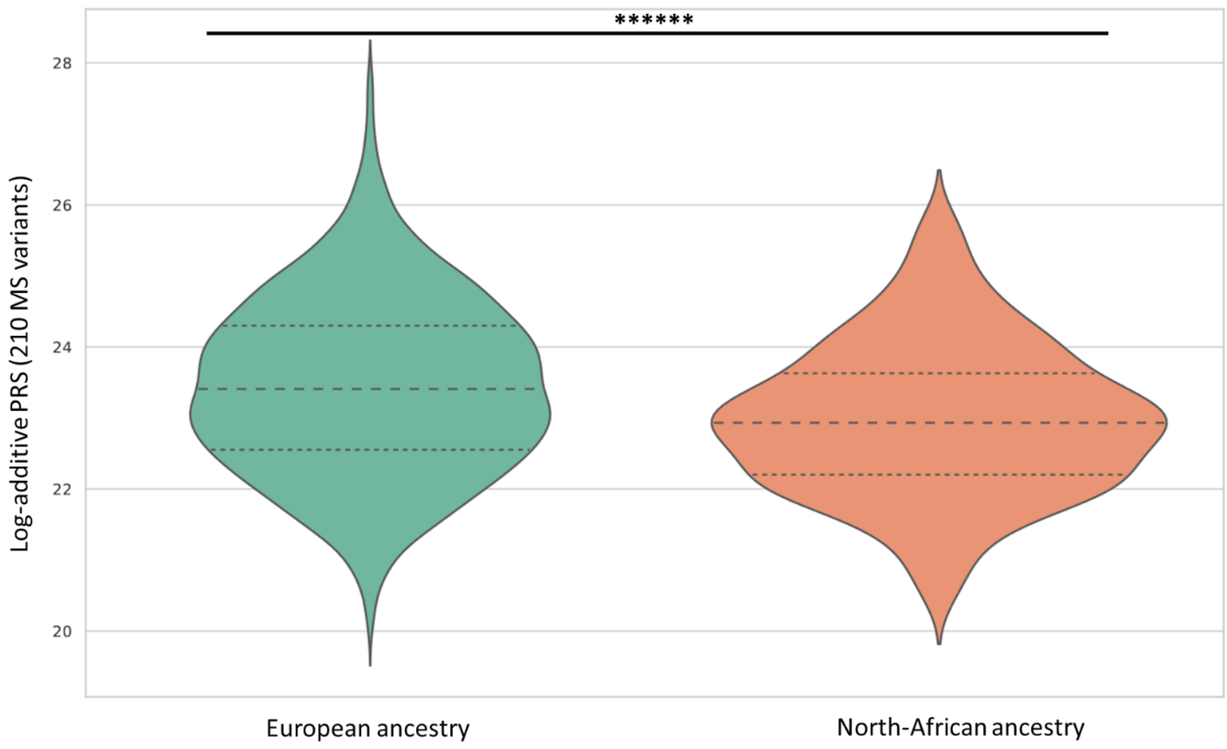


***Figure S22:*** *Distribution of log-additive PRS using all 210 available MS susceptibility variants, including MHC region variants. Non-parametric Mann-Whitney U test comparing European and North-African ancestry score distributions resulted in a statistically significant difference (p < 1e-6). Outer doted lines within violin plots represent 25th and 75th percentiles of the distributions. Inner doted lines represent the mean of the distributions.*
